# The case for using flexible healthcare capacity constraints to optimise pandemic control strategies

**DOI:** 10.1101/2025.10.09.25337668

**Authors:** Nathan J. Doyle, Fergus Cumming, Thomas J. R. Finnie, Michael West, Robin N. Thompson, Michael J. Tildesley

**Affiliations:** EPSRC Centre for Doctoral Training in Mathematics for Real-World Systems, Mathematics Institute, University of Warwick, Coventry, United Kingdom; The Zeeman Institute for Systems Biology & Infectious Disease Epidemiology Research, Mathematics Institute and School of Life Sciences, University of Warwick, Coventry, United Kingdom; Foreign, Commonwealth and Development Office, London, United Kingdom; UK Health Security Agency, London, United Kingdom; Modelling and Data-Science for Emergency Preparedness, Resilience, and Response, Chief Data Officer Group, UK Health Security Agency, Porton Down, United Kingdom; Mathematical Institute, University of Oxford, Oxford, United Kingdom

## Abstract

During the COVID-19 pandemic, a key challenge was designing control strategies that balance the benefits and costs of public health and social measures while preventing healthcare systems becoming overwhelmed. This was often implemented by epidemiological modellers as a binding (hard) constraint based on the maximum number of hospital beds that could be occupied at any given time. However, experience from the pandemic demonstrated that healthcare capacity depends not only on the number of beds available, but also on staffing and the availability of other resources. We argue that defining healthcare capacity using a single number (beds available) does not adequately capture pressures in healthcare settings as high occupancy is sustained. We therefore introduce a framework for implementing flexible (soft) constraints on healthcare capacity by allowing the cost of control strategies to depend continuously on intensive care unit (ICU) occupancy. We illustrate pandemic scenarios where using a soft constraint is better to capture long-term pressures on the healthcare system. Additionally, we highlight that explicitly accounting for uncertainty is essential to choose a robust control strategy. For optimal control studies to provide the best evidence to policy for future pandemics, they must carefully consider how their healthcare capacity constraints are implemented.

## 1 Introduction

A key concern at the onset of the COVID-19 pandemic was whether public health and social measures (PHSMs) would be sufficient to avoid healthcare system collapse [1]. Early modelling studies concluded that stringent interventions were required to suppress transmission, otherwise the peak healthcare demand would exceed the number of available intensive care unit (ICU) beds several times over [2–5]. Such studies were crucial in informing global policy responses [6]. For example, in the United Kingdom, modelling studies guided the response to the first wave in Spring 2020, leading to a disease control strategy designed to reduce and delay peak healthcare demand [2, 3, 7]. This delay in transmission allowed the National Health Service (NHS) valuable time to adapt capacity for exclusive COVID-19 treatment [8]. This involved various hospital provision interventions such as the use of private hospitals, the redeployment of staff and the cancellation of elective care [9]. Demand for COVID-19 treatment peaked on 12th April 2020, with 18,974 patients occupying hospital beds (2,881 of which occupied a bed equipped with mechanical ventilation) at national level [10]. The extent to which the epidemic curve needs to be “flattened” is linked to the capacity of the healthcare system [11].

A common interpretation of healthcare capacity is the number of hospital beds available in the system to treat patients presenting for care [12]. From November 2020 to June 2022, NHS England data indicated that there was a surplus of beds available to meet demand for COVID-19 treatment [13], adding to debate regarding whether the NHS was overwhelmed during the pandemic. Fong *et al*. have since argued that healthcare systems such as the NHS are complex institutions which “cannot be described through simple numerical counts of bed spaces, occupancy rates, equipment, and staff” [14]. Those authors highlight examples from the recent pandemic in which healthcare burden affected clinical outcomes. The procurement and training of additional staff lagged behind the rapid creation of ad hoc ICU beds set up in repurposed wards. This led to diluted staff-patient ratios exacerbated by staff absences due to COVID-19 illness. Such beds were often equipped with ventilators designed for use in anaesthesia, which are linked to worsening expected outcomes compared to ventilators designed for the treatment of critical respiratory disease [15]. Hospital occupancy also varied spatiotemporally. During the pandemic, there was a surplus of hospital beds nationally but occupancy occasionally exceeded the number of beds available at local (trust) level [16]. This led to a drastic increase in the transfer of critical care patients, which is known to increase mortality risk. In summary, healthcare capacity cannot be quantified straightforwardly in terms of the beds available in the system because this number does not wholly encapsulate the system’s capability to provide treatment. When the healthcare system undergoes significant stress – as NHS England did during the pandemic – patients can still be brought into hospitals for treatment when ICU beds have been exceeded; it is likely that they will, however, receive a diminished standard of care.

Modelling studies are useful for assessing the impact of PHSMs and generating evidence for effective outbreak response [17]. Optimal control studies in epidemiology (modelling studies which seek to optimise the use of interventions) often consider operational constraints in high-prevalence regimes [18]. Such studies often include such a constraint to preserve the healthcare system, typically using hospital beds as a proxy for capacity [19–21]. The optimisation problem features a binding (hard) constraint on the maximum number of hospital beds that can be occupied [22]. Control strategies that lead to a peak hospital occupancy that exceeds the number of beds available are rendered infeasible, and strategies with sustained periods of near-overwhelming occupancy are deemed feasible [21]. The latter strategy is particularly unsustainable over long time periods as ICU mortality risk increases as pressure on the healthcare system increases [23–25]. Ideally, such studies should include a mechanism which quantifies healthcare pressures beyond a bed count. Rather than introducing variables for staff and resources directly into the modelling framework – which can drastically increase model complexity and epistemic uncertainty – it can be useful to apply flexible (soft) constraints where periods of high healthcare strain are explicitly penalised regardless of whether healthcare demand exceeds beds available [26].

Pandemic control strategies rely on a clear definition for healthcare capacity, a quantity which was highly uncertain at the onset of the COVID-19 pandemic. For example, Ferguson *et al*. [2] estimated ICU surge capacity for Great Britain to be approximately 5,000 beds. This was later revised down to an estimate of 2,627 beds for England (England accounted for 87% of Great Britain’s population in 2020 [27]) by McCabe *et al*. [8]. A pandemic mitigation strategy which aims to maximise economic and social freedoms and reduce the non-disease harms of interventions is contingent on confidence that the healthcare system will not be overwhelmed. The strategy that is optimal under one assumed healthcare capacity may not be optimal under another assumed healthcare capacity. Since healthcare capacity may not be fully known in practice, it is necessary to account for such uncertainty in modelling studies.

In this paper, we show that the optimal control strategy is dependent upon different methods for considering healthcare capacity constraints. Specifically, we show that using a soft constraint for healthcare capacity can account for healthcare pressures beyond ICU bed occupancy [14]. We illustrate the general principle by considering a concise mathematical model that simulates emerging epidemics and tracks the number of individuals requiring hospital care. For each scenario we directly implement both hard and soft constraints on healthcare capacity by allowing the cost of hospitalisation per patient to vary with hospital occupancy over their length of stay. We highlight instances whereby the healthcare capacity constraint applied influences the inferred optimal policy decision. Furthermore, we show that it is essential to account for uncertainty in healthcare capacity when optimising control interventions. Without doing this, sub-optimal interventions may be identified when using a hard constraint on healthcare capacity.

## 2 Methods

### 2.1 Epidemiological Model

We consider the general scenario of an emerging novel outbreak in a setting where there is a fixed number of hospital beds available to treat patients requiring critical care due to severe disease. In this paper we are primarily interested in how the cost of an outbreak evolves at different levels of hospital occupancy, hence we deliberately keep our epidemiological model as flexible and transparent as possible to demonstrate proof of concept. The modelling framework incorporates pathogen transmission and disease progression of severe patients through ICU settings.

A renewal equation framework is used to generate pathogen transmission and allow for intervention changes to be mapped transparently into changes in the epidemic reproduction number. Renewal equations are widely used in epidemiological modelling, particularly for real-time inference of the reproduction number [28–30]. The model generates a time series of daily incidence, where the expected number of new symptomatic cases *I*(*t*) on day *t* is given by

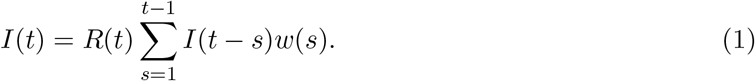

Here, *R*(*t*) denotes the instantaneous reproduction number: the average number of secondary cases that would arise from a primary case infected at calendar time *t* if conditions remained the same after time *t* [29]. The quantity *w*(*s*) represents the serial interval distribution: the time of symptom onset of a secondary case, measured from the time of symptom onset of the primary case *s*. The analagous serial interval distribution to the discrete probability distribution that we used is assumed to follow a Gamma distribution with mean 5.4 days and standard deviation 1.5 days, informed by Rai *et al*. [31]. Since the model framework produces discrete outputs we modify the Gamma distribution to obtain a discrete probability distribution for the serial interval *w* (see Supplementary Material).

The transmission model above (equation 1) is sufficient to compute new infected cases, but additional parameterisation is required to generate delayed quantities of interest relating to critical care outcomes. We extend the model to account for admissions and occupancy in ICU settings and generate corresponding time series for ICU incidence (ICU(*t*)) and prevalence (ICU^*o*^(*t*)). In this work we focus upon ICU occupancy rather than overall hospital occupancy given the stringency of that constraint during the COVID-19 pandemic [8] and the substantial negative health impact of losing access to critical care. In comparison, the constraint on the total number of hospital beds is less stringent, which we demonstrate by providing an analysis of outbreak dynamics for general hospital settings in the Supplementary Material (see Fig S2).

New ICU admissions on day *t* are computed using a parameter for the probability of requiring ICU care given symptoms *p*^ICU^ and a distribution for time from symptom onset to ICU admission *D*^I→ICU^(*t*)

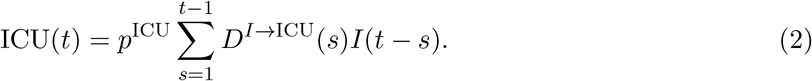

Patients are expected to spend a variable number of days in care; thus time-dependent quantities are required to compute occupancy in ICU settings. ICU occupancy 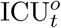 is dependent on *T* ^ICU^(*t*), the probability that a patient admitted to ICU still occupies a bed *t* days later.

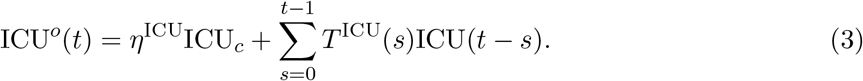

Here, we assume there exists a number of beds ICU_*c*_, of which a fraction *η*^ICU^ are occupied by emergency non-disease patients. We choose parameter values ICU_*c*_ = 4, 938 and *η*^ICU^ = 0.468 to match estimations of ICU bed occupancy across NHS England during the “surge phase” of the COVID-19 pandemic [8]. During this period (April-June 2020), measures were in place to reduce healthcare demand, such as the cancellation of elective care. The remaining beds available to treat patients admitted due to the outbreak is (1 *− η*^ICU^)ICU_*c*_. The epidemiological delay quantities *D*^*I→*ICU^(*t*), *T* ^ICU^(*t*) and the probability for clinical outcomes *p*^ICU^ are taken from the COVID-19 model developed by Keeling *et al*. [32], fitted to individual patient data during the first wave of the COVID-19 pandemic. As the source is an age-structured model, the parameters are aggregated by age (see Supplementary Material). Disease parameters and delay quantities are summarised in Table 1.

**Table 1.** Parameters in the disease and cost model.

| Parameter | Description | Value | Source |
| --- | --- | --- | --- |
| $p^{\text{ICU}}$ | Probability of ICU admission given symptoms | 0.0296 | [32] |
| $w$ | Serial interval distribution | Distribution | [31] |
| $D^{I \rightarrow \text{ICU}}$ | Time from symptoms to ICU | Distribution | [32] |
| $T^{\text{ICU}}$ | Probability a patient remains in ICU | Varies in $[0, 1]$ | [32] |
| $\text{ICU}_c$ | Number of hospital beds | 4,938 | [8] |
| $\eta^{\text{ICU}}$ | Background non-disease occupancy rate | 0.468 | [8] |
| $\alpha_0$ | Lower daily ICU cost value | 0.3 | [33] |
| $\alpha_1$ | Upper daily ICU cost value | 1 | [33] |
| $v$ | Rate of increase of cost | Varies | N/A |

### 2.2 Cost model

Here we present the framework for computing the health burden of the epidemic, dependent upon a policy maker’s healthcare capacity constraint. For simplicity, we measure the cost of the epidemic according to severe cases in ICU settings.

New ICU admissions on day *t* (ICU(*t*)) accumulate a cost which varies in response to changes in ICU occupancy over the patient’s length of stay, as occupancy can vary significantly at times of high epidemic growth or decline. We define *α* (*s*) as the daily cost per ICU case for future times *s* and weight it over the patient’s length of stay. The net cost of an outbreak is,

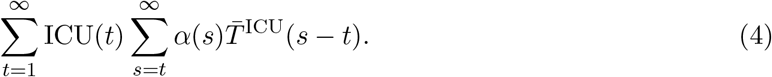

Here, 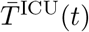 is the normalised probability that an individual is still occupying an ICU bed *t* days after admission, 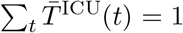 (Fig S1). For constant α(*t*) = 1, the total cost is exactly equal to the total number of ICU admissions. In this paper, we apply the constraint on healthcare capacity directly into the cost function by varying the daily cost per individual *α*(*t*).

#### 2.2.1 Applying constraints

The hard constraint is interpreted as a step increase in the cost variable *α*(*t*) once the total number of ICU beds ICU_*c*_ is exceeded by the number of patients requiring critical care due to severe disease (ICU(*t*)) or other emergency and elective care (*η*^ICU^ICU_*c*_). We choose a step function for *α* (*t*),

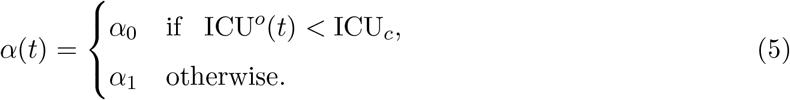

This step function has a lower value *α*_0_ representing a standard cost of ICU admission assuming a functioning healthcare system. The upper value *α*_1_ represents an elevated cost of ICU admission for a failed healthcare system. This can account for a range of adverse effects associated with the inability to adequately treat patients [14]. As an illustrative example, we set *α*_1_ = 1 and *α*_0_ = 0.3 based on the rough tripling of mortality rate due to severe COVID-19 illness in ICU settings [33].

For the soft constraint, we use a logistic formulation to smooth the step function (equation 5) around ICU_*c*_. This enables prevalence levels just below 100% occupancy to be penalised, accounting for worsening outcomes with a higher strain on ICU services. The time-dependent cost takes the following form,

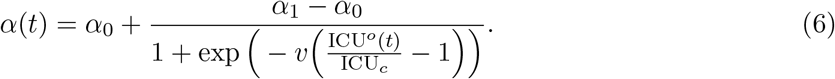

The parameter *v* controls the rate of increase of the logistic function from lower value *α*_0_ to upper value *α*_1_. This logistic function has an inflection point (α_0_ + α_1_)*/*2 at the point of 100% bed occupancy, ICU^*o*^(*t*) = ICU_*c*_. We chose such a smoothing function as it approaches the original step function, when *v → ∞*. Decreasing *v* increases smoothing and the severity of the constraint. By incorporating a soft constraint we intend to take two effects into account. As the healthcare system becomes saturated (but a small number of beds remain available), external factors such as staff, equipment and resources are stretched and the quality of care diminishes (increasing the cost per patient). Secondly, when demand for ICU beds exceeds supply the standard of care across the healthcare system is severely diminished but not every patient attains the elevated cost *α*_1_. Figure 1 demonstrates the behaviour of the cost α(*t*) and the difference between either constraint interpretation. We note that for *v <* 10, the soft constraint curve depicted in Fig 1 flattens such that the lower cost limit *α*_0_ is no longer attained as ICU(*t*) *→* 0. Costs are uniform regardless of occupancy level and this case no longer reflects the properties of a soft constraint, hence we restricted *v* to values greater than 10. Disease and cost parameters are outlined in Table 1.

**Figure 1:**
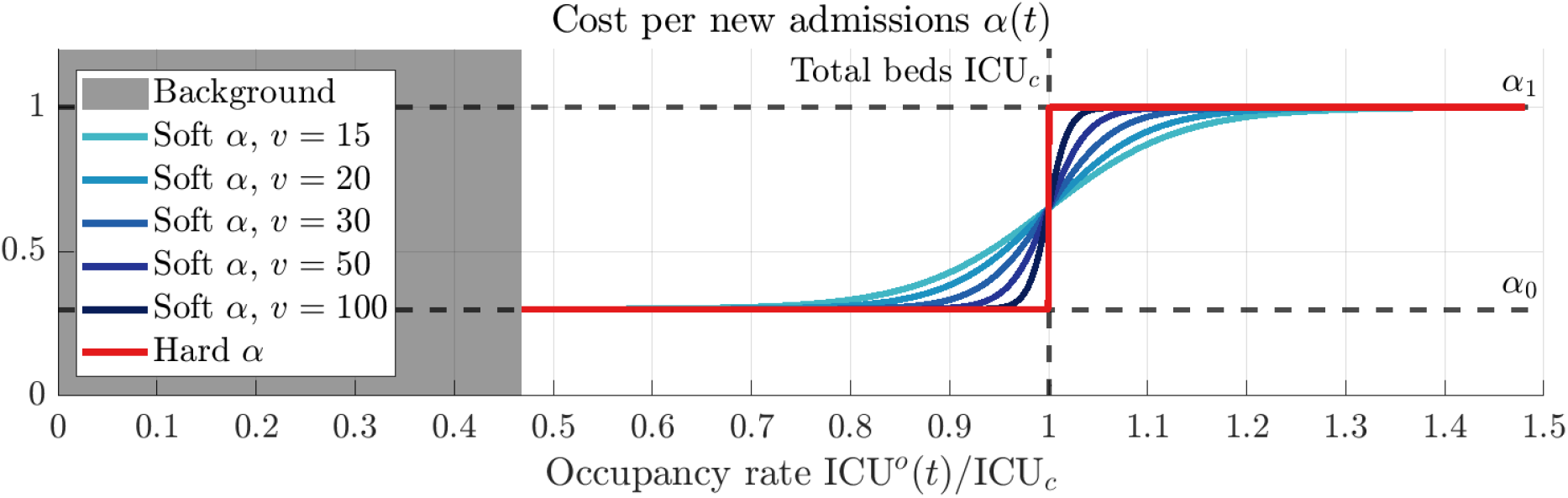
The time-dependent cost per ICU patient *α*(*t*) as a function of prevalence outlining the interpretation of either healthcare capacity constraint. The hard constraint (equation 5), highlighted in red increases sharply from the standard cost *α*_0_ to the elevated cost *α*_1_ once the total number of patients requiring critical care ICU^*o*^(*t*) surpasses the total number of beds ICU_*c*_. The soft constraint (equation 6), highlighted in blue smoothens the transition from *α*_0_ to *α*_1_. Decreasing the scaling factor *v* in the logistic function (moving from dark to light blue) increases the severity for which the soft constraint is applied. Shaded in grey is the assumed occupancy rate for non-disease patients *η*^ICU^ = 0.468.

#### 2.2.2 Incorporating uncertainty

We explore how variability in the true number of ICU beds, ICU_*c*_, and the soft constraint’s severity, *v*, could impact decision making. Estimating the relationship between the strain on healthcare services and ICU mortality risk can be challenging [34, 35] and may not be resolvable until after interventions have been implemented [8]. Given the novelty of the soft constraint approach, such uncertainties are of interest to a policy maker when evaluating options for disease mitigation. Hence, we consider scenarios whereby uncertainty exists at the outbreak onset and control strategies need to be assessed before uncertainty can be resolved.

We proceed by treating our variable of interest *x* = {*v, c* = (1 *− η*^ICU^)ICU_*c*_} as a random variable which follows a discretised normal distribution 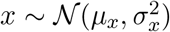 with mean *µ*_*x*_ and variance 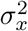. Details for the generation of a discretised probability mass function *f*_*x*_(*x*_*i*_) are provided in the Supplementary Material. We let *C*_*S*_ (*x*) be the function which computes the cost of strategy *S* using the soft constraint (equation 6) for uncertainty variable *x*. Since *C*_*S*_ (*x*) is a function of a random variable, we compute the expected cost of strategy *S* as follows,

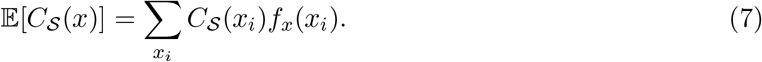

For the first analysis (Results 3.1), we treated the number of beds available (1 *− η*^ICU^)ICU_*c*_ as a random variable which was normally distributed *N* (*µ*_*c*_, *σ*^2^) with mean *µ*_*c*_ = 2, 627 and varying standard deviation *σ*_*c*_ *∈ {*100, 200, 300, 400*}* (see Fig S3 for distributions). In the face of uncertainty, we also compare outcomes at the 90th percentile of the outbreak cost distribution. This aims to reflect a policy maker’s desire to prepare for a “reasonable worst-case” scenario. Quantiles have previously been considered as cost statistics to minimise [36–38]. To reflect the preferences of a risk-averse policy maker, we compute the quantiles for each strategy cost distribution. For a quantile *q ∈* [0, 1], this is defined as the cost *Q*_*S*_ (*x*; *q*) such that P[*C*_*S*_ (*x*) *> Q*_*S*_ (*x*; *q*)] = 1 *− q*.

In the analysis presented, we use each metric to compare two mitigation strategies which are generated using the renewal model, equations (1)–(3).

## 3 Results

### 3.1 Scenario 1: Penalising high prevalence regimes

We identified scenarios where using a soft constraint for healthcare capacity – as opposed to a hard constraint – leads to different policy decisions. We first considered a scenario where policies that ensure low-prevalence can be optimal in cases where they appear sub-optimal when using a hard constraint on healthcare capacity. We used the model to simulate two outbreaks which differed in the amount of control applied, yet the peaks of neither exceeded the number of ICU beds available, ICU_*c*_. We first compared an “Early Suppression” (ES) strategy for disease control against a counterfactual “Late Suppression” (LS) strategy. For Strategy ES we used the renewal model, equations (1)-(3), to simulate an outbreak where the reproduction number took values *R*(*t*) = 2 for times *t ∈* [1, 63) and *R*(*t*) = 0.91 for times *t* ≥ 63 (Fig 2A). By selecting these values we assume the policy maker acts to reverse outbreak growth when the healthcare system is endangered. In comparison, for Strategy LS we set *R*(*t*) = 2 for times *t ∈* [1, 70) and *R*(*t*) = 0.7 for times *t ≥* 70. The *R*(*t*) values during interventions reflect the estimates for COVID-19 in the UK after lockdown was introduced in 2020 [39]. The policy maker’s intervention is delayed a week, but the delay allows the intervention to be more targeted (e.g., to high-risk groups) and is therefore more effective in reducing *R*(*t*). For this scenario, there is a trade-off between the time to enact the intervention and its effectiveness. Each outbreak was simulated for 750 days with initial condition *I*(1) = 10 symptomatic cases and ICU(1) = 0.

**Figure 2:**
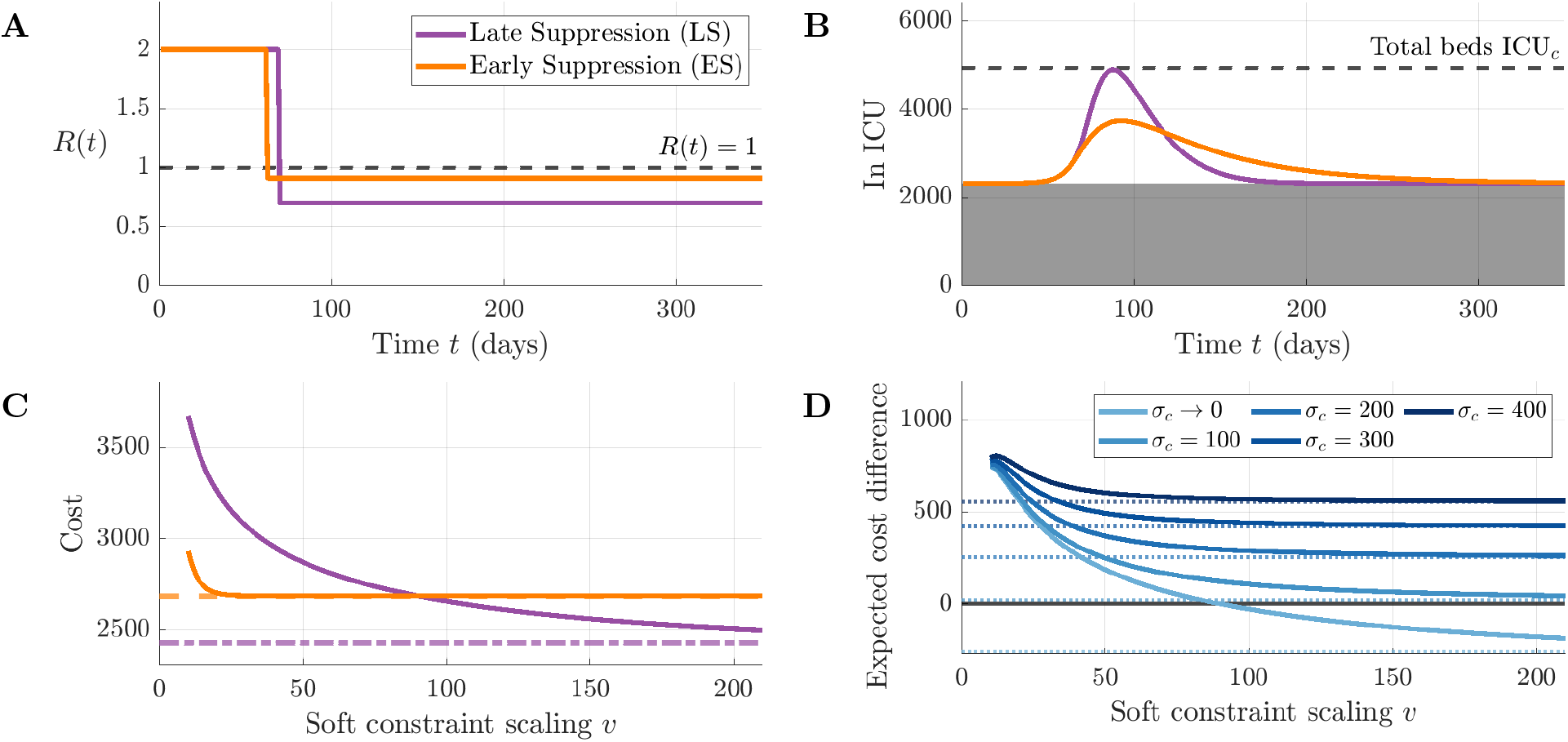
The simulation of the Late Suppression strategy (LS, purple) alongside the Early Suppression strategy (ES, orange) and their costs under the soft constraint. Each strategy is simulated from a time series for the reproduction number *R*(*t*) (A), leading to drastically different dynamics (B) The dashed black line indicates the total number of ICU beds available ICU_*c*_, and shaded in grey is the assumed occupancy level for non-disease patients. The optimal strategy (defined by lower cost) is dependent upon parameters *v* scaling the severity of the soft constraint (C). Horizontal lines mark the outbreak costs under the hard constraint. We also plot the expected benefit of Strategy ES, 𝔼[*C*_LS_(*c*)] *−* 𝔼[*C*_ES_(*c*)], subject to uncertainty in available ICU beds (D). A positive expected benefit corresponded to Strategy ES being optimal over Strategy LS. Dotted lines represent the expected benefit of Strategy ES under the hard constraint.

At our decision point *t* = 63 days into the outbreak, the strategies diverged. The earlier outbreak response adopted by Strategy ES acted to diminish and delay the timing of peak ICU prevalence (Fig 2B). While Strategy ES was successful in reversing outbreak growth, the epidemic took substantially longer to reach low levels of prevalence due to the effective reproduction number being marginally below one. In comparison, the delay to implement control in Strategy LS caused the ICU prevalence to dangerously approach (but not surpass) the number of available ICU beds Before the intervention at *t* = 70 days, disease prevalence in ICU doubled approximately every 7 days, equal to the intervention delay. However, since the intervention brought *R*(*t*) more substantially below one, the strategy returned to low prevalence levels quicker and had a lower total number of patients requiring ICU care as a result.

Both strategies were feasible when a hard constraint on healthcare capacity was used, and their cost depended on the final size of each outbreak (in terms of cumulative admissions to ICU). By this metric, Strategy ES was more expensive (the black dashed and dotted lines in Fig 2C). Under the soft constraint, Strategy LS is penalised for its high prevalence and Strategy ES became optimal for soft constraint scaling parameter *v <* 90.2 (comparing the orange and purple lines in Fig 2C).

Motivated by uncertainty in healthcare capacity in the UK early in the COVID-19 pandemic, we incorporated uncertainty in healthcare capacity to illustrate its impact on the optimal strategy. We vary the uncertainty in the availability of ICU beds by changing the parameter *σ*_*c*_, with larger values of *σ*_*c*_ corresponding to more uncertainty. In addition to the soft constraint parameterisation, the expected benefit of Strategy ES, 𝔼[*C*_LS_(*c*)] *−* 𝔼[*C*_ES_(*c*)], depended on uncertainty regarding available ICU beds (Fig 2D). More uncertainty (increasing *σ*_*c*_ lines in Fig 2D) led to an improved performance of Strategy ES compared to Strategy LS (defined as having lower expected cost for more values of *v* parameter space).

### 3.2 Scenario 2: Accounting for long-term strains on healthcare resources

In reality, briefly surpassing 100% bed occupancy might be expected to correspond to a lower outbreak cost in comparison to prolonged periods of strain on healthcare resources. The renewal model was used to simulate two outbreaks with interventions designed to reduce the reproduction number *R*(*t*) and mitigate transmission. We consider a “Running Hot” (RH) control strategy which arises as a solution in optimal control analyses [21, 40, 41]. This is pitted against a “Mistimed Suppression” (MS) strategy whereby the timing of a suppressive intervention was ill-judged and healthcare demand briefly surpasses beds available before the outbreak growth is reversed. Each outbreak was simulated for 750 days from the initial number of symptomatically infected cases *I*(1) = 10 and ICU(1) = 0.

Strategy RH was generated as an example scenario in which effectiveness dramatically differs between the two constraints considered. The reproduction number was set to *R*(*t*) = 2 for times *t ∈* [1, 64), *R*(*t*) = 1 for times *t ∈* [64, 214), and *R*(*t*) = 0.7 for times *t ≥* 214 (Fig 3A). For this strategy the policy maker is averse to economic and social harms, subject to maintaining a surplus of ICU beds in the system. After an initial growth phase, an intervention was implemented to halt but not reverse epidemic growth (Fig 3B). This resulted in a long period where ICU occupancy plateaued near but below the number of beds available to treat disease patients requiring critical care. Following this sustained period, the intervention was intensified to suppress transmission and return to low prevalence.

**Figure 3:**
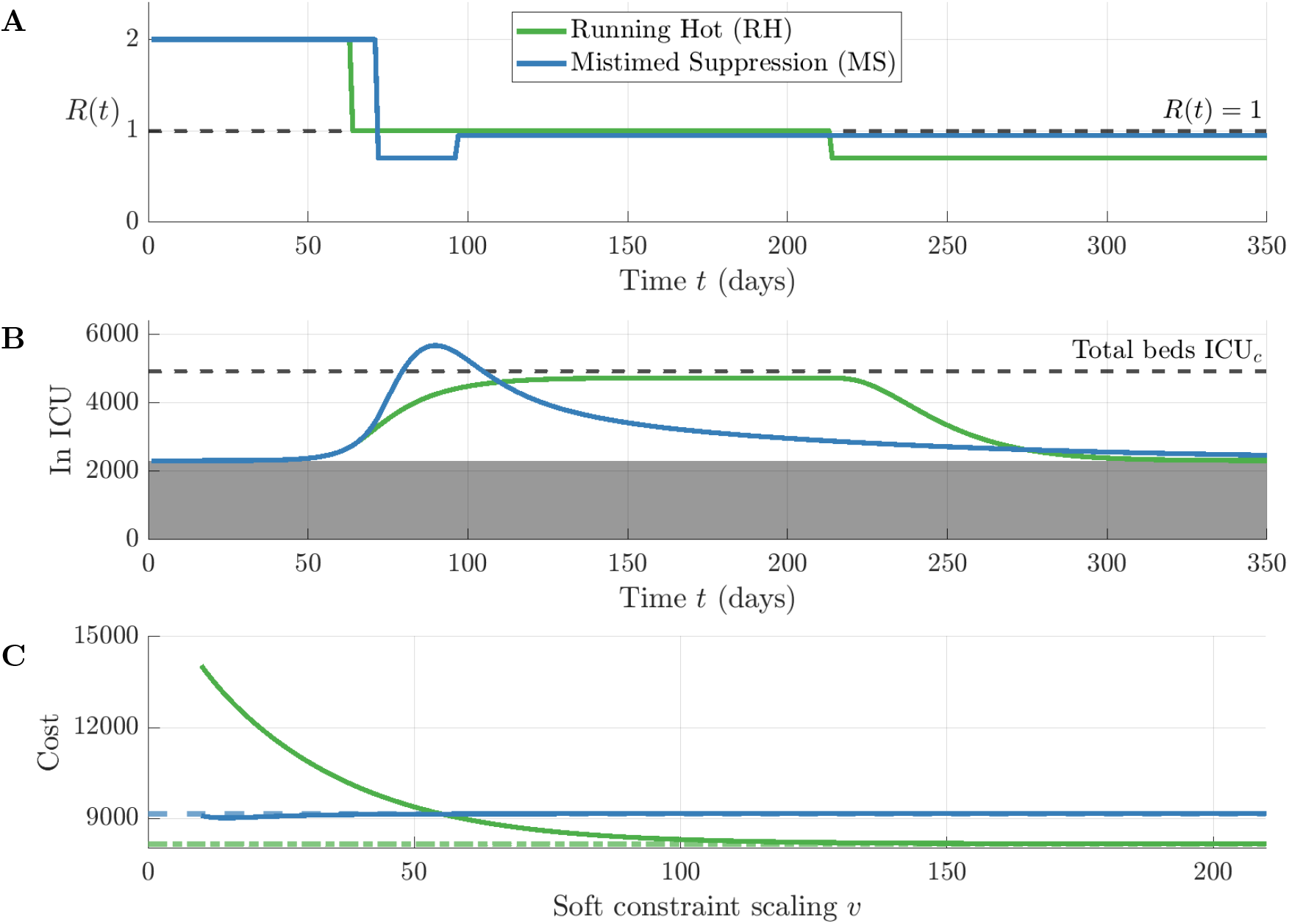
The simulation of the Running Hot strategy (RH, green) alongside the Mistimed Suppression strategy (MS, blue) and their costs under the soft constraint. Each strategy is simulated from a time series for the reproduction number *R*(*t*) (A), leading to drastically different dynamics (B). The dashed black line indicates the total number of ICU beds available ICU_*c*_. The optimal strategy (defined by lower cost) is dependent upon parameters *v* scaling the severity of the soft constraint (C). Horizontal lines mark the outbreak costs under the hard constraint. Note that cost is increasing on the y-axis and so the optimal strategy (defined by lower cost) lies below its counterpart. Shaded in grey is the assumed occupancy level for non-disease patients.

In comparison, Strategy MS was simulated to demonstrate the infeasibility of Strategy RH for the soft constraint framework (as might be expected in reality). The reproduction number was set to *R*(*t*) = 2 for times *t ∈* [1, 72), *R*(*t*) = 0.7 for times *t ∈* [72, 97), and *R*(*t*) = 0.95 for times *t ≥* 97 (Fig 3A). These (*t, R*(*t*)) values were chosen as an example where the policy maker wishes to suppress the outbreak, but fails to take delays in disease progression into account and thus mistimes the intervention. Measures to curb the initial growth phase were delayed slightly such that the peak ICU occupancy briefly exceeded the number of beds available (Fig 3B). There was a natural delay between changes in the transmission rate and the growth in ICU occupancy. The timing of peak ICU occupancy was delayed from the *R*(*t*) = 0.7 intervention onset as a result of the epidemiological delay quantities. After the peak, control measures suppressed the outbreak to low prevalence levels. This led to a lower total number of patients requiring ICU care (18,780 patients) compared to Strategy RH (27,172 patients).

When the costs of each strategy were computed using a hard constraint for healthcare capacity, Strategy MS was clearly deemed infeasible and was punished for surpassing the number of beds available (comparing the black dashed and dotted lines in Fig 3C). This was not necessarily true upon applying the soft constraint, where there was a transition at *v* = 55.2 below which Strategy MS became optimal over Strategy RH (comparing the blue and green lines in Fig 3C). Under the soft constraint, high ICU prevalence was penalised and the total cost of Strategy RH was higher for low values of the scaling factor *v*. In contrast, the total cost of Strategy MS did not vary substantially with *v*. The time period where ICU beds were exhausted was sufficiently brief such that its cost was balanced by the heightened costs for periods with near full occupancy. The decision boundary *v* = 55.2 corresponded to a logistic function which begins to noticeably diverge from the hard constraint framework when ICU bed occupancy exceeds 90-95% (Fig 1). For large *v* the cost of Strategy RH decreased as we approach the hard constraint framework (*v →∞*).

The results of this scenario demonstrate why Strategy RH is an expensive and undesirable strategy for disease mitigation, in comparison to a counterfactual (Strategy MS) which is already not a favourable policy for pandemic mitigation as it exceeds the number of beds available. The soft constraint framework was developed with the specific goal to penalise sustained periods of high stress on the healthcare system. Factors evident during the recent COVID-19 pandemic – such as staffing and regional pressures – lead to costs which may be difficult to model explicitly, but should be considered in the evaluation of control strategies.

### 3.3 Uncertainty in soft constraint severity

By treating the scaling factor in the soft constraint *v* as uncertain (see Fig 4A for sample distributions), we considered the full cost distribution of each strategy. The nature of each control strategy led to distributions with fundamentally different shapes (Fig 4B). The cost distribution for Strategy MS was narrow since the majority of the outbreak was spent at low prevalence levels with only a brief window of time with ICU demand close to the number of beds available. This resulted in a net cost which was largely independent of the severity of the soft constraint *v*. In contrast, Strategy RH was associated with a long-tailed distribution and was not robust to uncertainty in *v*. For large *v* we approach the hard constraint and the effects of strain on healthcare services near 100% occupancy are neglected and costs were lower. However, net costs increased sharply as *v* decreased. The shapes of these cost distributions did not vary qualitatively with changes in the mean *µ*_*v*_ (Fig S5).

**Figure 4:**
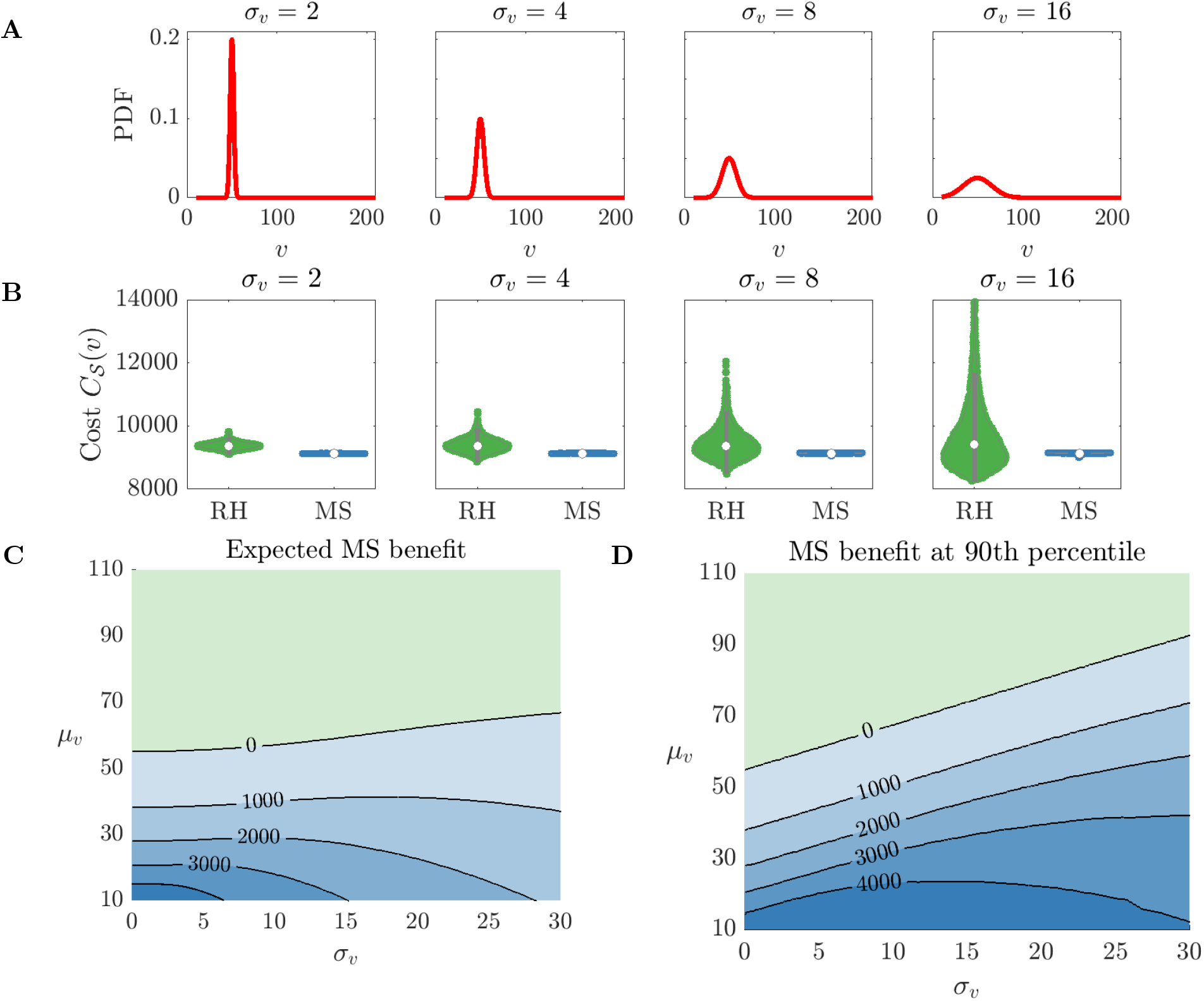
We investigate uncertainty in applying the soft constraint by treating the soft constraint parameter *v* as a normal random variable 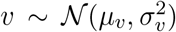 with *µ*_*v*_ = 50 and varying *σ*_*v*_ (A). A distribution in the underlying soft constraint yields cost distributions for each strategy (B). For varying *µ*_*v*_, *σ*_*v*_ characterising our soft constraint distribution we use the resulting cost distributions to compute the benefit of Strategy MS with regards to expectation 𝔼[*C*_RH_(*v*)] *−* 𝔼[*C*_MS_(*v*)] (C) and the 90th percentile of the strategy cost distributions *𝒬*_RH_(*v*; 0.9) *− 𝒬*_MS_(*v*; 0.9) (D). A positive benefit corresponded to Strategy MS being optimal over Strategy RH. Blue indicates parameter regimes where Strategy MS is optimal and green indicates areas where Strategy RH is optimal. Violin plots were generated using Violinplot-Matlab [42].

We calculated the expected cost of each strategy 𝔼[*C*_*S*_ (*v*)] using equation (7). The expected benefit of Strategy MS 𝔼[*C*_RH_(*v*)] *−* 𝔼[*C*_MS_(*v*)] varied with the distribution’s mean *µ*_*v*_ to a greater extent in comparison to its standard deviation *σ*_*v*_ (Fig 4C). The decision boundary (the line in (*σ*_*v*_, *µ*_*v*_) space where 𝔼[*C*_RH_(*v*)] = 𝔼[*C*_MS_(*v*)] increased with *σ*_*v*_. This corresponded to an increase in the area in (*µ*_*v*_, *σ*_*v*_) parameter space under which MS had the lower expected cost. Since the total cost varied for Strategy RH more significantly in comparison to Strategy MS, increasing the width of the *v* distribution led to a greater frequency in high cost outcomes for Strategy RH. For very large uncertainty *σ*_*v*_ *>>* 1, the expected benefit of Strategy MS diminished as the *v* distribution flattens. We also computed the 90th percentile of the strategy cost distributions *Q*_RH_(*v*; 0.9) *− Q*_MS_(*v*; 0.9) (Fig 4D). As expected from Fig 4B, a higher quantile *q* = 0.9 to minimise against made Strategy MS optimal over Strategy RH for a larger area of (*µ*_*v*_, *σ*_*v*_) parameter space. Furthermore, the decision boundary (the line in (*σ*_*v*_, *µ*_*v*_) space where the optimal strategy switched) increased with *σ*_*v*_. This indicates that Strategy MS appears more robust to a risk-averse policy maker as uncertainty in the soft constraint’s severity is considered.

Finally we demonstrate that strategies which ensure high prevalence, such as Strategy RH, require an exact estimation of available ICU beds to treat patients. We assume the number of non-disease emergency patients *η*^ICU^ICU_*c*_ was known and we compute the cost of each strategy as (1 *− η*^ICU^)ICU_*c*_ is varied from 1,000 to 4,000. It became apparent that Strategy RH was only optimal when certainty in the number of available ICU beds was high (Fig S4B). This highlights a key failure of strategies that aim to maintain demand for healthcare services just below healthcare capacity since they rely on exact estimates of wider healthcare capacity to ensure the system does not get overwhelmed. Given that uncertainty, such strategies could underestimate the degree of mitigation required to maintain a functioning healthcare system.

## 4 Discussion

A resilient healthcare system is essential for an effective pandemic response. Modelling studies which project short-term healthcare demand are increasingly used to assess the impact of control measures and guide policy decisions [2–4, 43]. However, the metric for healthcare demand is typically hospital and/or ICU beds occupied in relation to a (fixed) number of beds available in the healthcare system. The baseline number of beds available to treat patients is just one component of a wider system, the quality of care can vary substantially with healthcare demand, even if the number of beds required is not exceeded. This effect is overlooked when using ICU bed occupancy as a hard constraint to identify optimal strategies for pandemic management.

We introduce a simple cost framework for implementing hard and soft constraints on healthcare capacity for optimal disease control problems. The soft constraint framework is designed to use the number of beds as a proxy for capacity, but with a cost per new patient that increases with the strain on healthcare services. We used this framework to outline cases where the inferred optimal control strategy changed with the class of healthcare constraint applied. We first investigated a decision problem where the policy maker was faced with two interventions of different timings and intensities (Strategies LS, ES). While neither strategy led to the number of concurrent ICU patients exceeding the number of beds available, earlier action to reverse epidemic growth (Strategy ES) was more likely to be optimal under a soft constraint and considerations for uncertainty regarding the number of available ICU beds. We then investigated a scenario where briefly surpassing ICU beds available (Strategy MS) was less costly in some instances compared to a prolonged period marginally below 100% ICU bed occupancy (Strategy RH). Strategies similar to Strategy RH are often favoured in mathematical optimal control studies based on hard constraints, yet in practice they bring an unsustainable burden on the healthcare system. The soft constraint framework was conceptualised to better represent healthcare capacity, recognising that it is not entirely captured by the 100% bed occupancy threshold, and to explore the implications for optimal control strategies. Further, we have shown that soft constraints allow for the identification of strategies which are more robust to uncertainty in ICU bed provision. The cost distributions for Strategies MS and RH were vastly different in width, and the latter strategy would be unlikely to be preferable to a risk-averse policy maker. This also provides additional motivation for policy makers to resolve uncertainties surrounding outbreak costs under differing levels of strain on the healthcare system. If these uncertainties can be reduced, then more effective control measures may be identified.

This work serves as an important contribution to optimal control research in epidemiology. Many optimal control studies specific to the COVID-19 pandemic implemented healthcare capacity constraints that were hard by design [19–22]. Sensitivity to its value can be tested [19, 21], however in each scenario it remains a hard constraint and the qualitative behaviour of optimal strategies does not change. In particular, this can lead to solutions which deploy just enough control to ensure bed capacity is not exceeded. In practice, these policies are unsustainable as the wider system faces pressures which can decrease the efficacy of care. Soft constraints on healthcare capacity have been applied in a limited number of settings. Kantner *et al*. [21] employed a soft constraint by allowing the probability of severe clinical outcomes to increase drastically once bed resources are exhausted. However, in that study, this effect is not captured at levels just below 100% occupancy, so ICU prevalence in optimal solutions ends up straddling this critical boundary. Our soft constraint framework is similar to the work of Schnyder *et al*. [26] who allowed the cost of infection to vary as a function of prevalence such that it increases dramatically as healthcare settings become saturated. We build on their work in a number of areas, and undertake a direct comparison of inferred optimal interventions using soft and hard constraints in different scenarios. Additionally, we explicitly explore uncertainty in its severity to highlight the (lack of) robustness of certain mitigation strategies. Other literature has explored the use of “tier” structures for which PHSMs are intensified and relaxed with prevalence [44–47], as was implemented in the UK in October–December 2020 during the COVID-19 pandemic. If these were to be implemented in a future pandemic, penalising interventions that are more likely to lead to prevalence necessitating the implementation of higher tiers, and therefore a higher strain on healthcare services, is a consideration that we stress in this work.

Our modelling framework is deliberately kept simple to highlight the key result that the use of soft constraints on healthcare capacity can lead to different control strategies that are more likely to be optimal in the real-world. The model can be extended to incorporate additional realism, such as heterogeneity in transmission between individuals of different ages [48], at the expense of additional parameter uncertainty. More detailed models exist for determining the allocation of hospital interventions when resources are limited [49, 50]. Our soft constraint framework can be applied to such cases to aid with the design of optimal interventions when there is a high strain on healthcare services. The trajectory of any simulated outbreak is subject to assumptions about the effectiveness of interventions. We assume background occupancy *η*^ICU^ remains constant across all scenarios for simplicity, though seasonal variations are possible [51]. Despite our modelling simplifications, we would expect the importance of using a soft constraint to reflect the strain on healthcare services to hold irrespective of the complexity of the underlying epidemiological model and our soft constraint framework can be applied during outbreaks of a range of pathogens with high pandemic potential. We demonstrate that the key messages of our paper are robust to changes in model parameterisation. We present that the message holds in both scenarios when the timing and intensity of interventions differ for Strategies ES and MS (See Supplemental Material). The uncertainty in costing is explored in the main text by treating *v* as a random variable, in addition to sensitivity analysis for the upper and lower cost values to ICU admission *α*_0_ and *α*_1_ (Fig S6-S9). We note that our cost framework is applicable in scenarios similar to COVID-19 where policy makers sought to manage a large-scale pandemic when healthcare systems were at risk of saturation. In the early stages of an outbreak, policy makers may consider the possibility of eliminating the outbreak and in such cases a different modelling framework or cost function may be more applicable. When containment has failed and elimination becomes extremely costly, policy makers must balance trade-offs such as the costs of interventions and the benefits of maintaining low numbers of cases [52]. Background treatment in our model (captured by parameter *η*^ICU^) was informed by an estimation of the impact of cancelling elective care in the United Kingdom during the surge phase of the COVID-19 pandemic [8]. The cancellation of elective care can have devastating long-term health impacts [53], particularly in lower income groups [54]. Capturing such indirect costs is beyond the scope of this study, but is an additional consideration important to policy makers.

A limitation of many outbreak optimal control analyses lies in the difficulty in resolving the relationship between ICU occupancy and mortality in real-time during an outbreak. The literature which quantifies this causation in the context of the COVID-19 pandemic has always been retrospectively determined [23, 24, 34, 35]. This motivated us to explore uncertainty in this relationship by varying the degree to which we apply the soft constraint quantified by parameter *v*. For future pandemics, it can be useful to base this parameterisation from established thresholds for “safe occupancy” which vary between 85-92% ICU bed occupancy for the NHS [16, 55]. These 85% and 92% thresholds equate to substantial costs when the soft constraint is parameterised for *v <* 25 and *v <* 75, respectively (Figure 1). Understanding how the strain on healthcare services affects mortality, and working where possible to resolve uncertainties surrounding this relationship, should be a key consideration for future pandemic preparedness initiatives. This study motivates a range of important considerations for future work. Healthcare occupancy can vary seasonally [51], which would influence the class of optimal control strategies in periods where the healthcare system is coping with winter pressures. Healthcare capacity (and the capability to adapt capacity in a crisis) can vary significantly between countries [56]. Thus, the class of mitigation strategies which are optimal in one setting may not be viable in another. As hospital occupancy varied spatiotemporally during the COVID-19 pandemic, intervention strategies must preserve capacity at regional level rather than at the national scale to minimise the use of critical care transfers. The identification of such strategies – which would require a spatial modelling framework – is a key area for future work. For policy makers to design effective control strategies during future pandemics, they must first define their objectives for optimal disease management [38, 45]. If a key goal is to ensure that healthcare services do not become overstretched, then we contend that using a soft constraint to represent the pressure on healthcare services, rather than a hard constraint, is most appropriate. As we have shown, a soft constraint properly reflects the fact that case numbers being maintained just below physical capacity for a long period is unlikely to be sustainable. Constraints should account for a range of different pressures on healthcare services (e.g. additional staffing requirements driven by staff sickness) in addition to bed counts [14]. Inferred optimal control strategies under soft constraints tend to be more robust to epidemiological uncertainties, since they do not rely on knowing precisely when hospital capacity has been reached. We hope that our research motivates the use of soft constraints in future epidemiological cost-benefit analyses, enabling policy makers to optimise control interventions more effectively during future outbreaks.

## Data Availability

All data generated or analysed during this study, including computing code for reproducing our results, are available at: https://github.com/ndoyle815/soft-constraints-hospital-capacity-v1?tab=readme-ov-file

## Author contributions

**Nathan J. Doyle:** Conceptualization, Formal analysis, Investigation, Methodology, Validation, Visualization, Writing - original draft, Writing – review & editing. **Fergus Cumming:** Conceptualization, Writing – review & editing. **Thomas J. R. Finnie:** Conceptualization, Writing – review & editing. **Michael West:** Conceptualization, Writing – review & editing. **Robin N. Thompson:** Conceptualization, Methodology, Supervision, Writing – review & editing. **Michael J. Tildesley:** Conceptualization, Funding acquisition, Methodology, Supervision, Writing – review & editing.

## Competing interests

We declare that we have no competing interest.

## Data and code availability

All data generated or analysed during this study, including computing code for reproducing our results, are available at: https://github.com/ndoyle815/soft-constraints-hospital-capacity-vtab=readme-ov-file.

## Funding

N.J.D. and M.J.T. were supported by the Engineering and Physical Sciences Research Council through the Mathematics of Systems II Centre for Doctoral Training at the University of Warwick (grant number EP/S022244/1). The funders had no role in study design, data collection and analysis, decision to publish, or preparation of the manuscript.

## Notes

### Competing Interest Statement

The authors have declared no competing interest.

### Summary of Updates

Author affiliations updated, supplementary file uploaded

