## Supplementary Material for "The case for using flexible healthcare capacity constraints to optimise pandemic control strategies"

#### Contents

**1 Model parameters and distributions** **2**

**2 Generating uncertainty** **3**

**3 Dynamics in General and Acute hospital settings** **4**

**4 Uncertainty in ICU beds available** **5**

**5 Scenario 2: Long-tailed cost distribution for RH** **6**

**6 Sensitivity to lower and upper ICU cost values** **7**

**7 Sensitivity to the timing and effectiveness of interventions** **13**

**8 Results with discounting costs at 1.5% per annum** **15**

#### 1 Model parameters and distributions

The serial interval for the disease is assumed to follow a Gamma distribution with mean 5.4 days and standard deviation 1.5 days [1]. Since the renewal equation framework requires a discrete probability distribution, we discretised this continuous distribution (with probability density function $\bar{w}(t), t \in [0, \infty)$) to obtain a discrete serial interval distribution (with probability mass function $w(s), s = \{1, 2, \ldots\}$). We applied a method previously used by Cori *et al.* [2],

$$w(s) = \frac{1}{\Omega} \int_{s-1}^{s+1} (1 - |u - s|)\bar{w}(u)du, \tag{1}$$

where $\Omega$ is a normalisation constant to ensure that $w$ represents a valid probability mass function. The epidemiological delay quantities relating to ICU admissions, namely the time from symptom onset to ICU admission $D^{I \to \mathrm{ICU}}$ and the time spent in ICU $T^{\mathrm{ICU}}$, were sourced from Keeling *et al.* [3] and were already discretised for daily increments and are presented in Fig S1.

Three disease parameters are derived from an age-structured model [3], namely the probability of eventual ICU admission given symptoms $p$, the delay distribution from symptom onset to ICU admission $D^{I \to H}$ and the quantity characterising the ICU length of stay $T^H$. To apply the original age-specific quantities to this study, they were aggregated from the original five-year age bands $G' = \{0-4, 5-9, \ldots, 95-100, 100+\}$. The aggregated quantity of interest $q \in \{p, D^{I \to H}, T^H\}$ for our model is computed as follows,

$$q = \sum_{g \in G'} N'_g q'_g, \tag{2}$$

where $N'_g$ is the proportion of the total United Kingdom population which reside in age group $g$ (also taken from [3]) and $q'_g$ is the original age-structured quantity for age bands $G'$.

As an illustrative example, we parameterise the lower value of daily cost per ICU admission $\alpha_0$ by the mortality rate in ICU settings. We set $\alpha_1 = 1$ so that cost is measured in units of $\alpha_1$ (the number of ICU patients who lose provision of adequate critical care). If we were to express these costs in terms of Quality Adjusted Life Years (QALYs), the ratio $\alpha_0/\alpha_1$ does not significantly change. This is because the average QALY loss due to COVID-19 death (accurred over years of life lost) is significantly greater than QALY losses due to non-fatal ICU admission (accurred over short-term acute care). For reference, Zala *et al.* [4] estimated the average QALY loss per COVID-19 death at 8.79882 and the average QALY loss per bed day at 0.0011 (ICU) and 0.00002 (non-ICU).

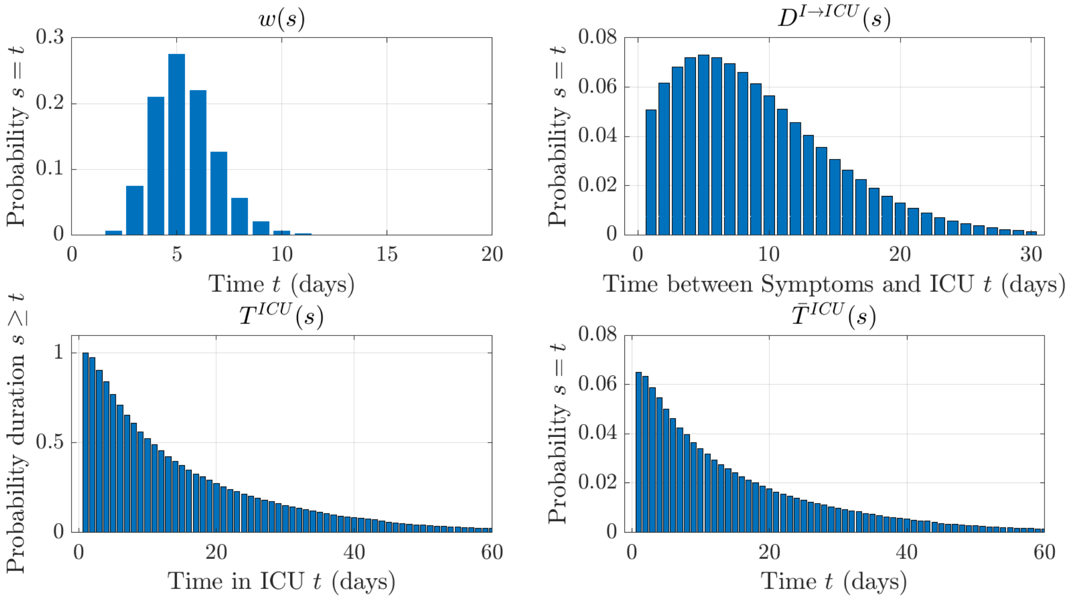


**Figure S1:** Model distributions and delay quantities, showing the discrete serial interval distribution $w$, the time from symptom onset to ICU admission $D^{I\to\text{ICU}}$, the quantity characterising the ICU length of stay (expressed as a daily probability of remaining in critical care $T^{\text{ICU}}$ and as a weighting in the cost model $\bar{T}^{\text{ICU}}$). The serial interval distribution was discretised from a Gamma distribution with mean 5.4 days and standard deviation 1.5 days, informed by Rai *et al.* [1]. The distribution $D^{I\to\text{ICU}}$ and quantity $T^{\text{ICU}}$ were sourced and aggregated from the COVID-19 model developed by Keeling *et al.* [3].

#### 2 Generating uncertainty

In our analysis we consider the effects of uncertainty in a random variable $x = \{v, c = (1 - \eta^{\text{ICU}})\text{ICU}_c\}$. For simplicity, we consider scenarios where our uncertainty variable is normally distributed $x \sim \mathcal{N}(\mu_x, \sigma_x^2)$ with mean $\mu_x$ and variance $\sigma_x^2$. For each $(\mu_x, \sigma_x)$ pair on support $[x_1, x_2]$, we generate the discretised probability mass function,

$$f_x(x_i) = \mathbb{P}[x = x_i] = \begin{cases} \mathbb{1}_{x_i=\mu_x} & \sigma_x = 0, \\ \int_{-\infty}^{x_i} \frac{1}{\sigma_x \Omega_x} \exp\left(\frac{(\xi-\mu_x)^2}{2\sigma_x^2}\right) d\xi & x = x_1, \sigma_x \neq 0, \\ \int_{x_i}^{\infty} \frac{1}{\sigma_x \Omega_x} \exp\left(\frac{(\xi-\mu_x)^2}{2\sigma_x^2}\right) d\xi & x = x_2, \sigma_x \neq 0, \\ \frac{1}{\sigma_x \Omega_x} \exp\left(\frac{(x_i-\mu_x)^2}{2\sigma_x^2}\right) & \text{otherwise}, \end{cases} \quad (3)$$

where $\Omega_x$ is a normalisation constant to ensure a valid probability mass function ($\sum_{x_i} f_x(x_i) = 1$).

When we investigate uncertainty in the soft constraint's severity $v$, we allow $\mu_v$ to take values in $[10, 210]$ with increments of 0.2, and we allow the standard deviation $\sigma_v$ to take values in $[0, 50]$

with increments of 0.25. Uncertainty in ICU beds available is incorporated by fixing the number of non-disease ICU patients $\eta^{\text{ICU}}\text{ICU}_c$ and varying the number of available ICU beds $(1-\eta^{\text{ICU}})\text{ICU}_c$ with $\mu_c = 2,627$ fixed and $\sigma_c \in \{100, 200, 300, 400\}$ (see Fig S3).

### 3 Dynamics in General and Acute hospital settings

In our main analysis we explore how potential causalities between ICU strain and mortality risk impacts the effectiveness of mitigation control strategies. We chose to limit our costing of outbreaks to ICU settings since the ICU bed constraint was significantly more stringent compared to the general hospital bed constraint during the COVID-19 pandemic [5]. Since time-dependent interventions are implemented in our analysis in response to the saturation of ICU beds, we do not approach the risk of exceeding the number of general hospital beds available. We demonstrate this by extending the renewal modelling framework to track the number of individuals requiring access to general and acute hospital care.

New general (excluding ICU) hospital admissions $H(t)$ on day $t$ arise from a proportion $p^H$ of infected cases who eventually require general hospital care given symptomatic infection. The delay between symptom onset and hospitalisation is characterised by the distribution $D^{I\to H}(t)$,

$$H(t) = p^H \sum_{s=1}^{t-1} D^{I\to H}(s) I(t-s). \tag{4}$$

To compute occupancy for general hospital beds $H^o(t)$, we require a quantity $T^H(t)$ , the probability that a patient admitted to a non-ICU hospital bed still occupies a bed $t$ days later. Critical care patients are typically admitted to a bed on a normal ward following time in ICU, and so we include a similar quantity $T^{\text{ICU}\to H}(t)$ for the time an ICU patient occupies a hospital bed on a normal ward after discharge from critical care.

$$H^o(t) = \eta^H H_c + \sum_{s=0}^{t-1} T^H(s) H(t-s) + \sum_{s=0}^{t-1} T^{\text{ICU}\to H}(s)\text{ICU}(t-s). \tag{5}$$

The disease parameters and distributions $p^H, D^{I\to H}, T^H, T^{H\to\text{ICU}}$ derive from the Warwick COVID-19 model [3] and are also age-aggregated using the method described in 1. Background hospital occupancy and beds available are sourced from bed limit definitions during the surge phase of the COVID-19 pandemic. [5]. Specifically, we simulated hospital dynamics concurrently for the Running Hot (RH) and Mistimed Suppression (MS) strategies with $p^H = 0.1441, \eta^H = 0.4597, H_c = 11,525$. For both strategies, the timing and intensity of interventions was a response to ICU occupancy rates which curbed outbreak growth (Fig S2). Evidently, the lack of ICU beds constrained the maximum prevalence of the outbreak and we never threatened to surpass the total number of general hospital beds $H_c$.

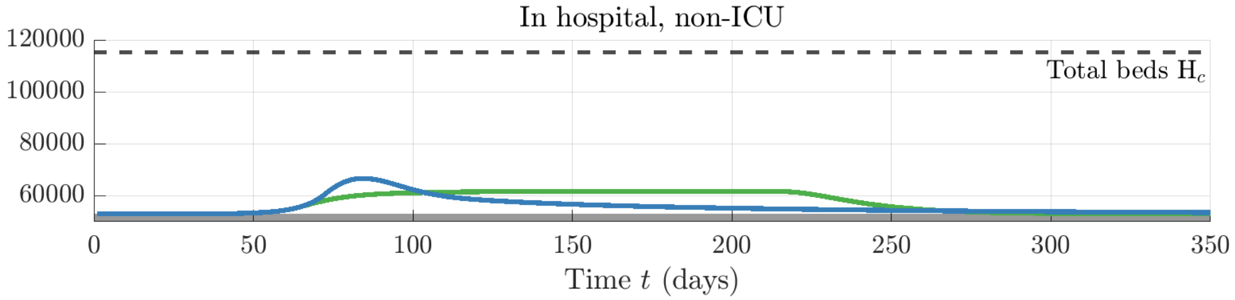


**Figure S2:** Outbreak dynamics in general hospital settings for the Running Hot strategy (RH, green) and the Mistimed Suppression strategy (MS, blue) using the equations (4)-(5) extended to the renewal framework. Background non-disease occupancy in general hospital settings $\eta^H H_c$ is shaded in grey.

### 4 Uncertainty in ICU beds available

In the main analysis (Fig 2D), we modelled available ICU beds as normally distributed $(1 - \eta^{\text{ICU}})\text{ICU}_c \sim \mathcal{N}(\mu_c, \sigma_c)$ with $\mu_c = 2,627$ and $\sigma_c \in 100, 200, 300, 400$. These distributions correspond to the following 95% credible intervals: $[2,431,\ 2,823]$ for $\sigma = 100$, $[2,235,\ 3,019]$ for $\sigma = 200$, $[2,039,\ 3,215]$ for $\sigma = 300$ and $[1,843,\ 3,409]$ for $\sigma = 400$. For reference, we present these distributions below.

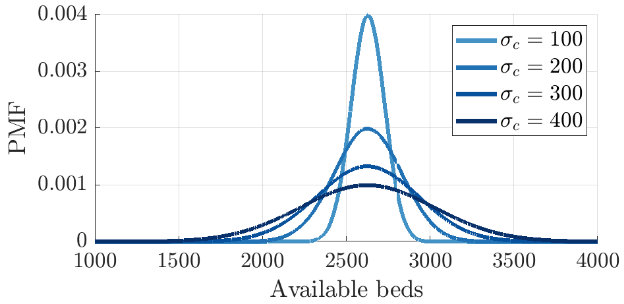


**Figure S3:** Distributions for the number of available ICU beds $(1 - \eta^{\text{ICU}})\text{ICU}_c \sim \mathcal{N}(\mu_c, \sigma_c)$ used to produce Fig 2D in the main analysis.

We demonstrate how the performance of Late Suppression (Strategy LS) against Early Suppression (Strategy ES) varies with uncertainty in the number of available ICU beds $(1 - \eta^{\text{ICU}})\text{ICU}_c$. For varying $v \in [10, 210]$ and $(1 - \eta^{\text{ICU}})\text{ICU}_c \in [1,000,\ 4,000]$ we computed the difference in costs between Strategies LS and ES. We repeat this analysis for the second scenario to compare the Running Hot (RH) and Mistimed Suppression (MS) control strategies, computing the difference in costs between Strategies RH and MS (Fig S4B). We show that the available number of ICU beds has an overwhelming effect on the performance of each control strategy, and their efficacy requires a precise estimation of healthcare bed constraints.

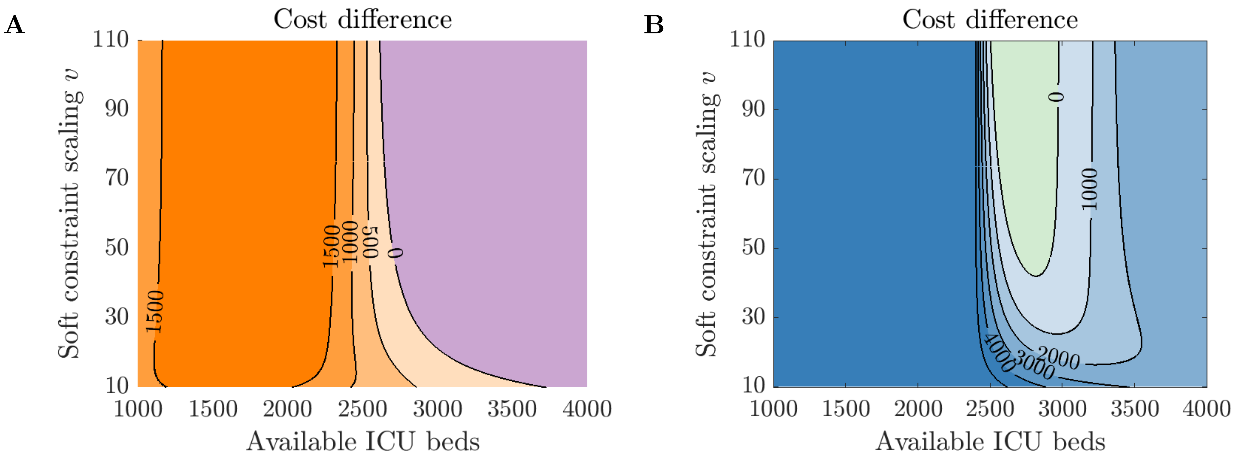


**Figure S4:** Difference in costs of the Late Suppression (LS, purple) and the Early Suppression (ES, blue) control strategies (A), in addition to the Difference in costs of the Running Hot (RH, green) and the Mistimed Suppression (MS, blue) control strategies (B), for varying the severity of the soft constraint $v$ and the number of available ICU beds $(1-\eta^{\mathrm{ICU}})\mathrm{ICU}_c$. For each panel, a positive cost difference corresponds to Strategy ES being optimal over Strategy LS, and Strategy MS being optimal over Strategy RH.

#### 5 Scenario 2: Long-tailed cost distribution for RH

Here we present the cost distributions for the Running Hot (RH) and Mistimed Suppression (MS) strategies as a result of the mean scaling of the soft constraint $\mu_v$. In the main analysis we presented results with $\mu_v = 50$ and showed that Strategy RH fundamentally has a long-tailed distribution. We show that this is invariant of higher $\mu_v$ where the expected cost of Strategy RH is lower, $\mathbb{E}[C_{\mathrm{RH}}(v)] < \mathbb{E}[C_{\mathrm{MS}}(v)]$.

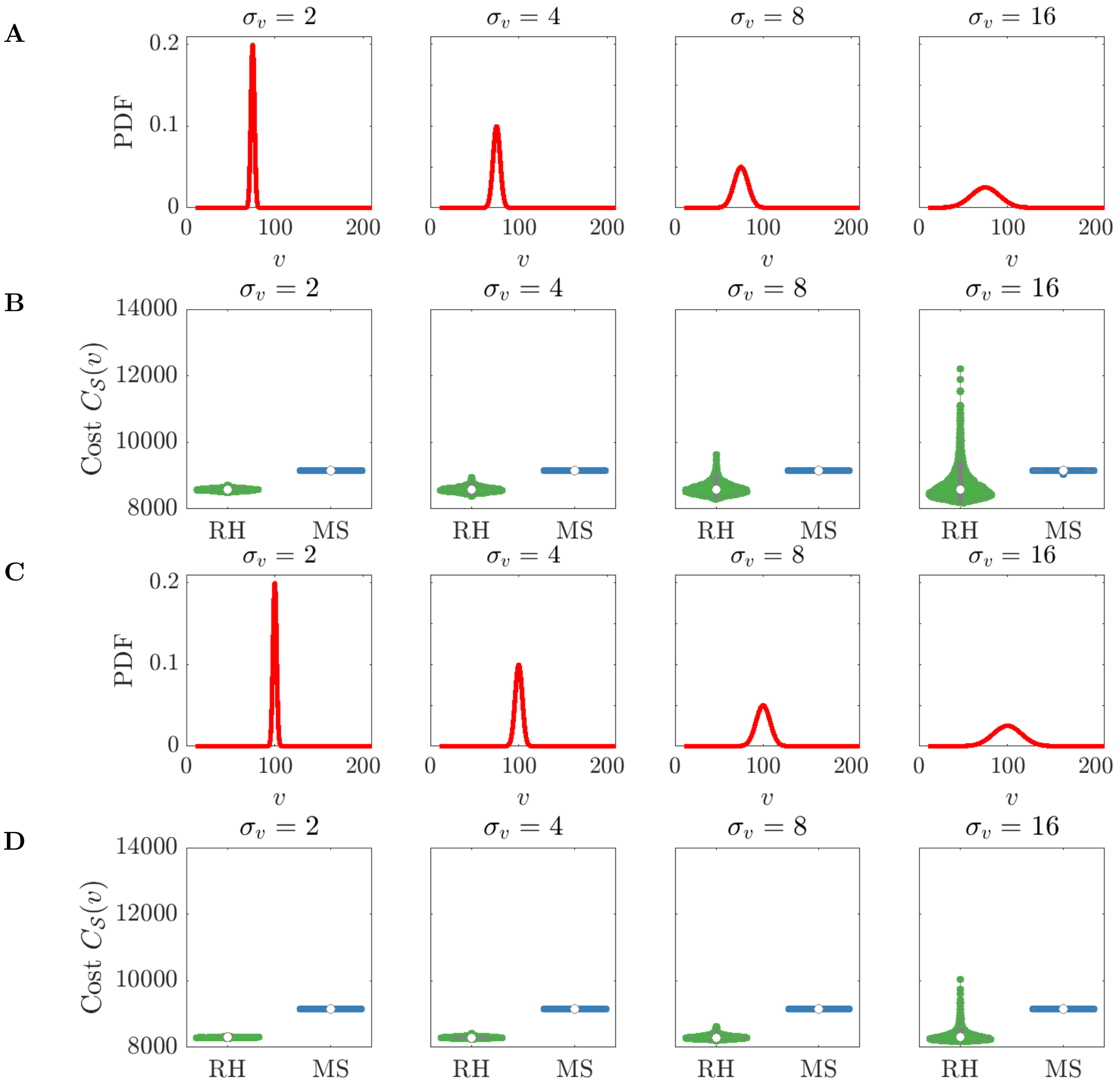


**Figure S5:** A distribution for parameterising the soft constraint $v \sim \mathcal{N}(\mu_v, \sigma_v)$ (A,C) yields cost distributions for each control strategy (B,D). The mean of the soft constraint distribution is increased to $\mu_v = 75$ (A,B) and $\mu_v = 100$ (C,D) to represent instances where the Running Hot strategy (RH, green) is optimal over the Mistimed Suppression strategy (MS, blue) under expectation, $\mathbb{E}[C_{\mathrm{RH}}(v)] < \mathbb{E}[C_{\mathrm{MS}}(v)]$.

#### 6 Sensitivity to lower and upper ICU cost values

We demonstrate the results are robust to changes in the standard cost for ICU admission, $\alpha_0$. Decreasing $\alpha_0$ can represent changes which lead to improved treatment in ICU setting, such as antivirals or a vaccine. We present examples with lower ($\alpha_0 = 0.2$) and greater ($\alpha_0 = 0.5$) baseline costs to ICU admission, with $\alpha_1 = 1$ fixed. For the elevated case $\alpha_0 = 0.5$ exceeding ICU beds available has a less significant effect on the health burden, yet the soft constraint still implies similar strategy outcomes (Figure S6A). In Figures S8 - S9, the discrepancy in outbreak final sizes between strategies RH and MS means their relative costs scale differently subject to changes in $\alpha_0$. However, when the timings of the $R(t) = 0.7$ intervention for Strategy MS are allowed to differ slightly we obtain a similar epidemic curve which reaches the same conclusions as the main text. High prevalence control strategies (such as the Running Hot strategy) need a soft constraint mechanism to demonstrate the negative impact of long-term healthcare strain. Poor strategies which exceed ICU beds available can be lower in cost under the soft constraint, demonstrating the failure of Strategy RH. Similar conclusions are drawn from allowing the elevated cost $\alpha_1$ to vary ($\alpha_1 = 5\alpha_0 = 1.5$ and $\alpha_1 = 2\alpha_0 = 0.6$), with $\alpha_0$ fixed (Figures S7A, S10, S11). Increased $\alpha_1$ may represent additional indirect consequences for exceeding ICU bed capacity which a policy maker might consider (e.g., further cancellations in elective care procedures), which we have not captured in our cost function to keep the framework tractable.

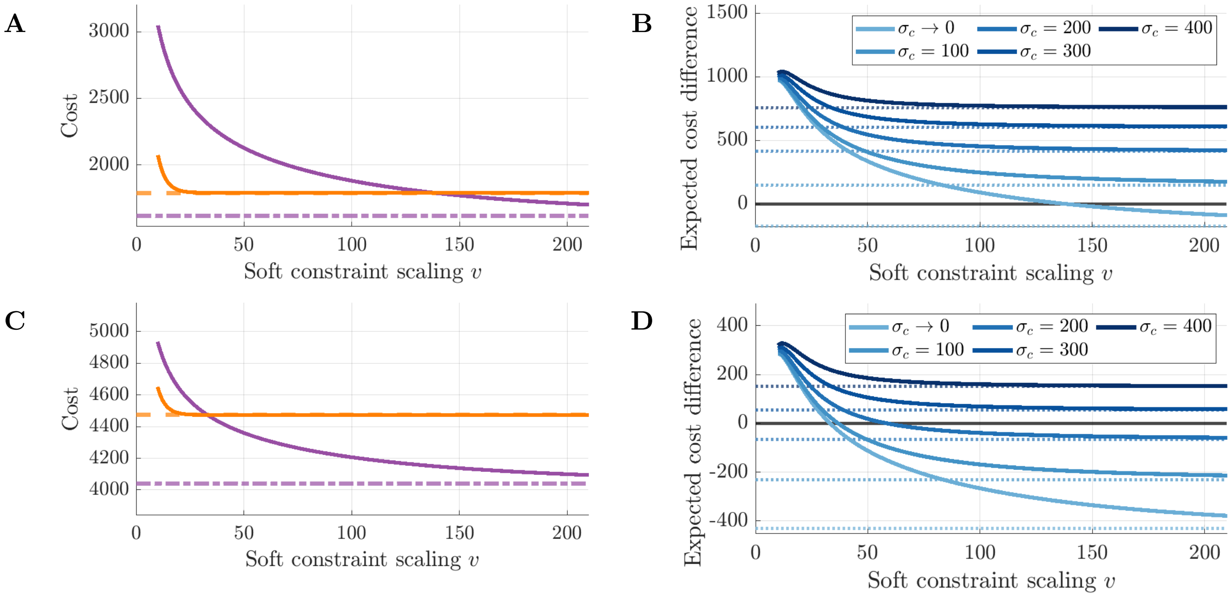


**Figure S6:** Plotting the optimal strategy (defined by the lower cost) between the Early Suppression strategy (Strategy ES, orange) and the Late Suppression strategy (Strategy LS, purple) under the soft constraint for $\alpha_0 = 0.2$ (A) and $\alpha_0 = 0.5$ (C). We use a black lines (dashed for Strategy ES, dotted for Strategy LS) to mark the outbreak costs under the hard constraint. Also presented is the difference in expected costs $\mathbb{E}[C_{\text{LS}}(v)] = \mathbb{E}[C_{\text{ES}}(v)]$ subject to uncertainty regarding the available number of ICU beds for $\alpha_0 = 0.2$ (B) and $\alpha_0 = 0.5$ (D). A positive difference in expected costs corresponds to Strategy ES being optimal over Strategy LS. Dotted lines represent the difference in expected costs under the hard constraint.

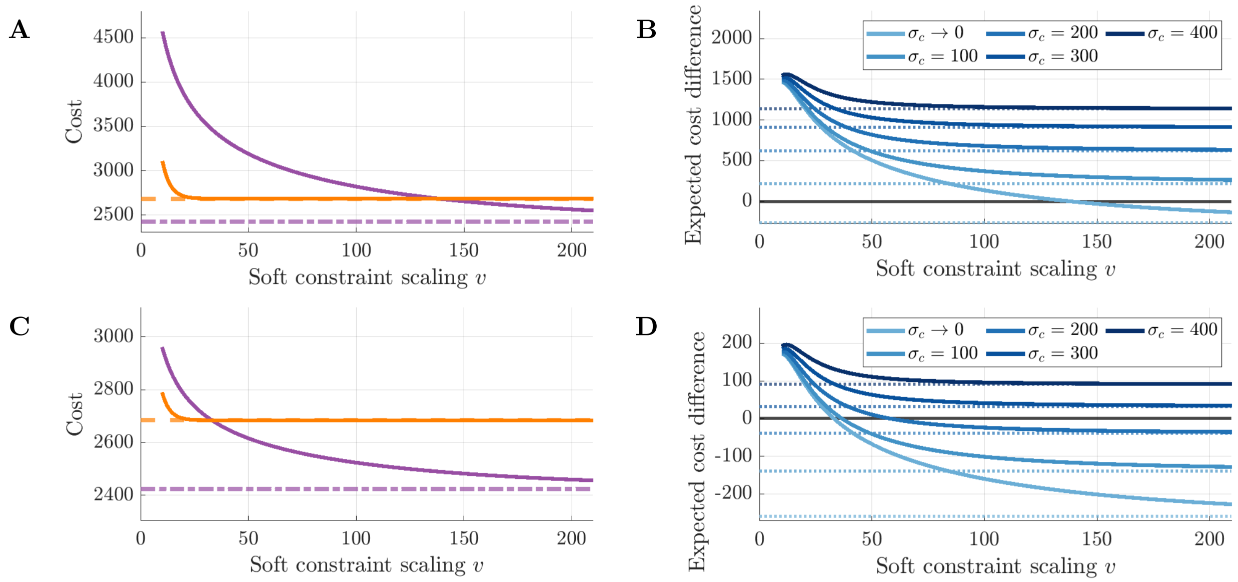


**Figure S7:** Plotting the optimal strategy (defined by the lower cost) between the Early Suppression strategy (Strategy ES, orange) and the Late Suppression strategy (Strategy LS, purple) under the soft constraint for $\alpha_1 = 5\alpha_0 = 1.5$ (A) and $\alpha_1 = 2\alpha_0 = 0.6$ (C). We use a black lines (dashed for Strategy ES, dotted for Strategy LS) to mark the outbreak costs under the hard constraint. Also presented is the difference in expected costs $\mathbb{E}[C_{\mathrm{LS}}(v)] = \mathbb{E}[C_{\mathrm{ES}}(v)]$ subject to uncertainty regarding the available number of ICU beds for $\alpha_0 = 1.5$ (B) and $\alpha_0 = 0.6$ (D). A positive difference in expected costs corresponds to Strategy ES being optimal over Strategy LS. Dotted lines represent the difference in expected costs under the hard constraint.

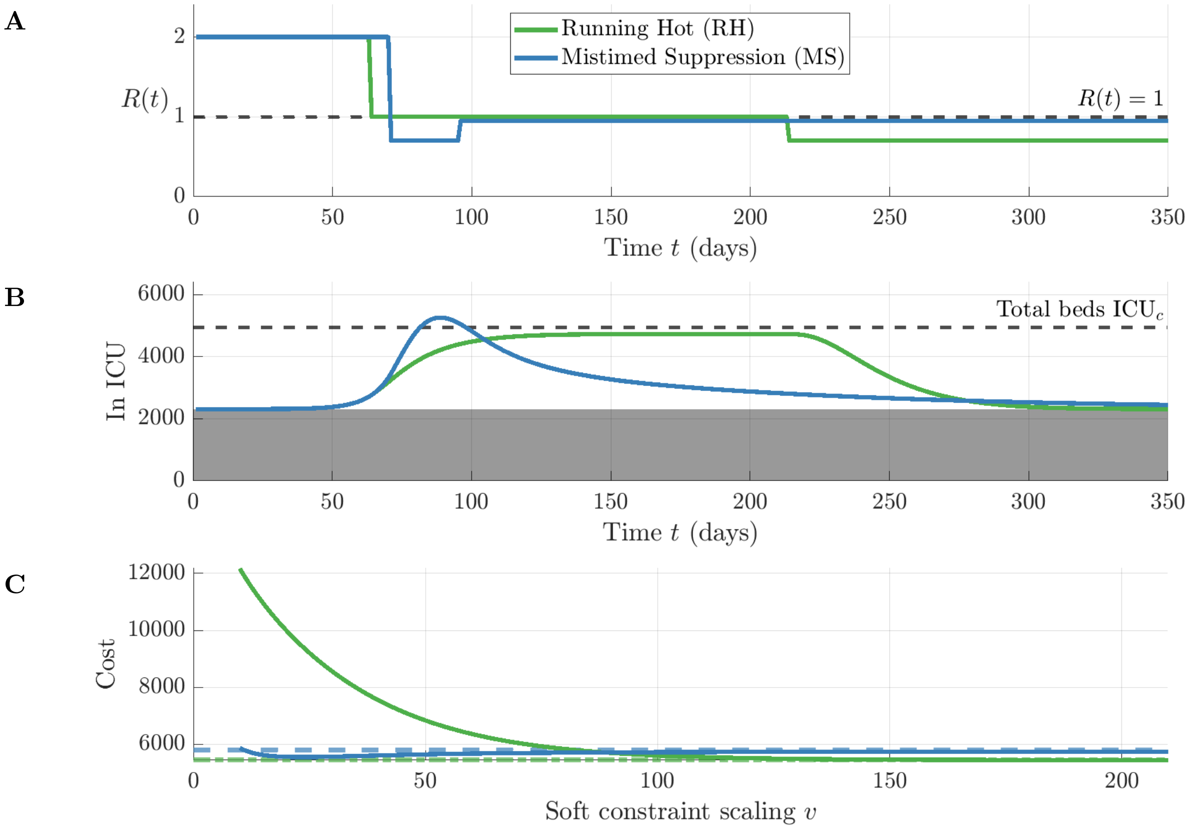


**Figure S8:** The simulation of the Running Hot strategy (RH, green) alongside the Mistimed Suppression strategy (MS, blue) and their costs under the soft constraint and $\alpha_0 = 0.2$. Each strategy is simulated from a time series for the reproduction number $R(t)$ (A), leading to drastically different dynamics (B). Here, the timing of the $R(t) = 0.7$ intervention for Strategy MS is one day earlier (day 71) in comparison to the main text (day 72). The dashed black line indicates capacity to treat disease patients $(1 - \eta^{\text{ICU}})\text{ICU}_c$. The optimal strategy (defined by lower cost) is dependent upon parameters $v$ scaling the severity of the soft constraint (C). Note that cost is increasing on the y-axis and so the optimal strategy (defined by lower cost) lies below its counterpart.

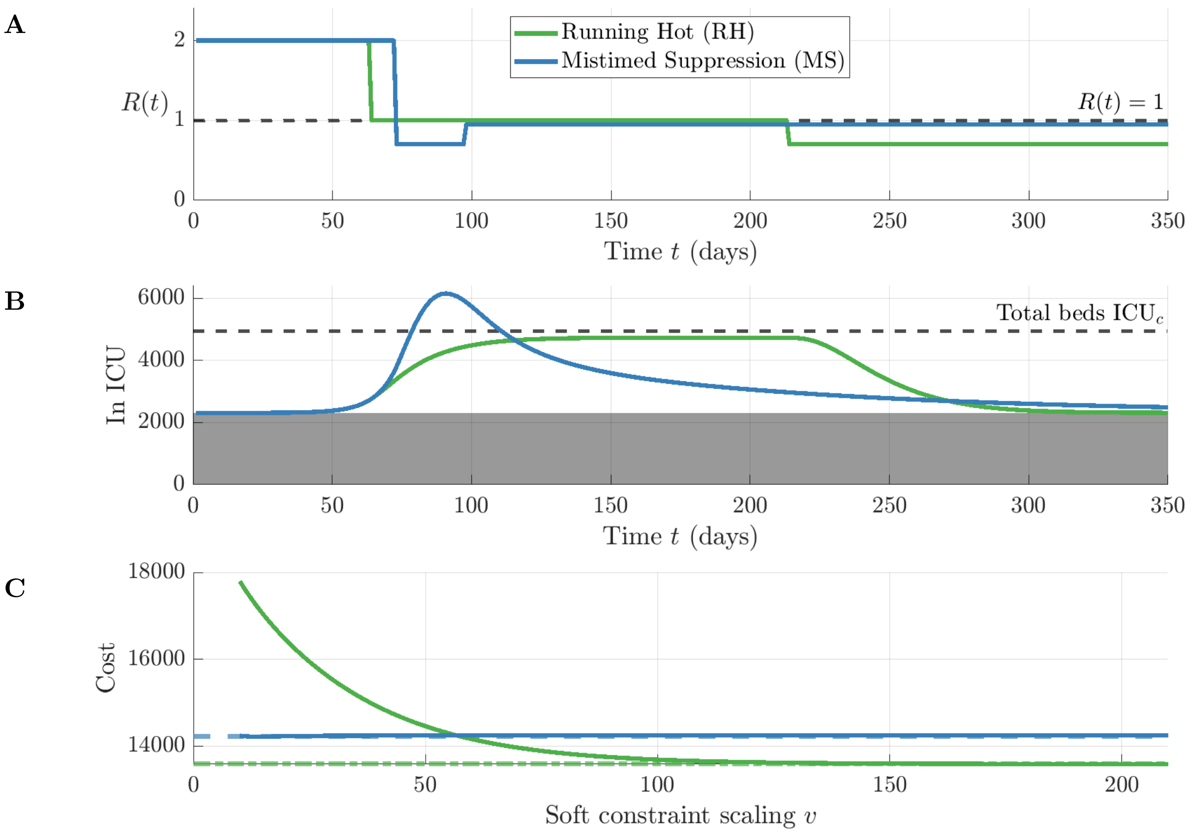


**Figure S9:** The simulation of the Running Hot strategy (RH, green) alongside the Mistimed Suppression strategy (MS, blue) and their costs under the soft constraint and $\alpha_0 = 0.5$. Each strategy is simulated from a time series for the reproduction number $R(t)$ (A), leading to drastically different dynamics (B). Here, the timing of the $R(t) = 0.7$ intervention for Strategy MS is one day later (day 73) in comparison to the main text (day 72). The dashed black line indicates capacity to treat disease patients $(1 - \eta^{\text{ICU}})\text{ICU}_c$. The optimal strategy (defined by lower cost) is dependent upon parameters $v$ scaling the severity of the soft constraint (C). Note that cost is increasing on the y-axis and so the optimal strategy (defined by lower cost) lies below its counterpart.

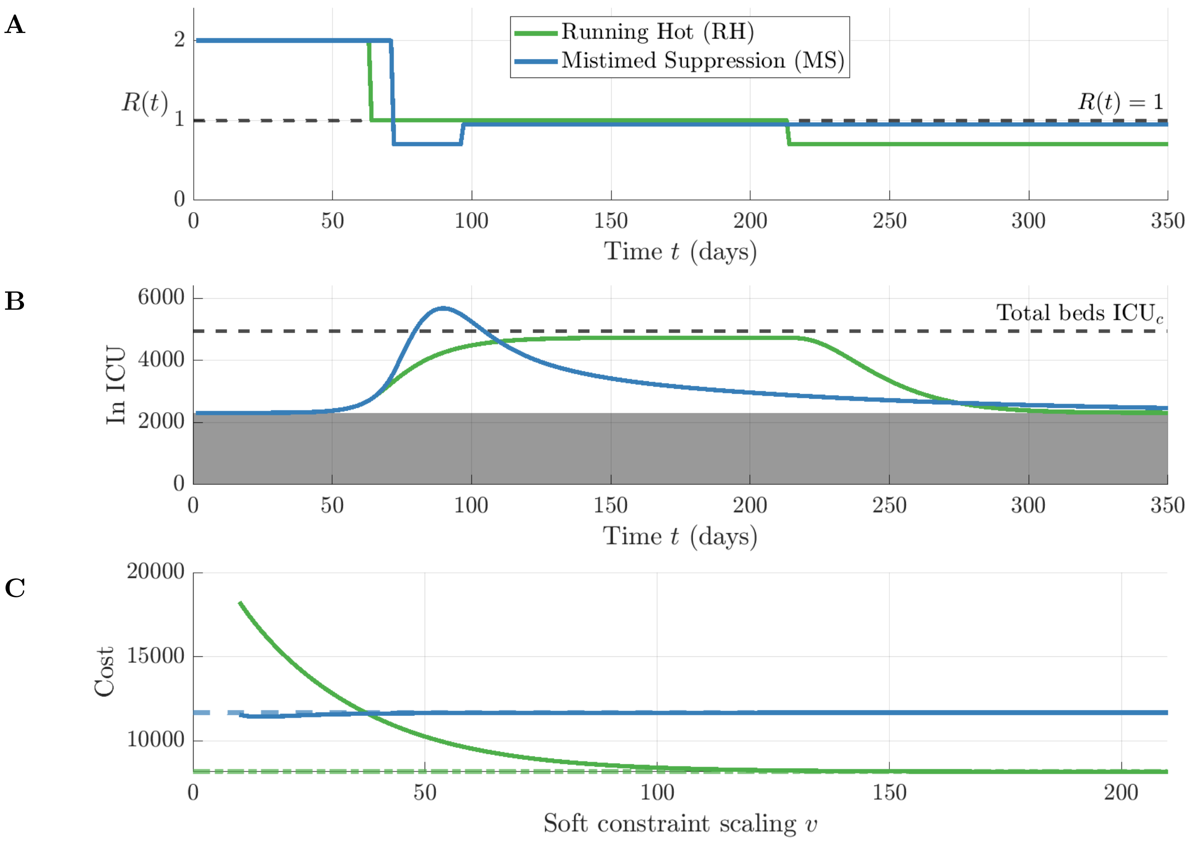


**Figure S10:** The simulation of the Running Hot strategy (RH, green) alongside the Mistimed Suppression strategy (MS, blue) and their costs under the soft constraint and $\alpha_1 = 2\alpha_0 = 1.5$. Each strategy is simulated from a time series for the reproduction number $R(t)$ (A), leading to drastically different dynamics (B). Here, the timing of the $R(t) = 0.7$ intervention for Strategy MS is one day later (day 73) in comparison to the main text (day 72). The dashed black line indicates capacity to treat disease patients $(1 - \eta^{\mathrm{ICU}})\mathrm{ICU}_c$. The optimal strategy (defined by lower cost) is dependent upon parameters $v$ scaling the severity of the soft constraint (C). Note that cost is increasing on the y-axis and so the optimal strategy (defined by lower cost) lies below its counterpart.

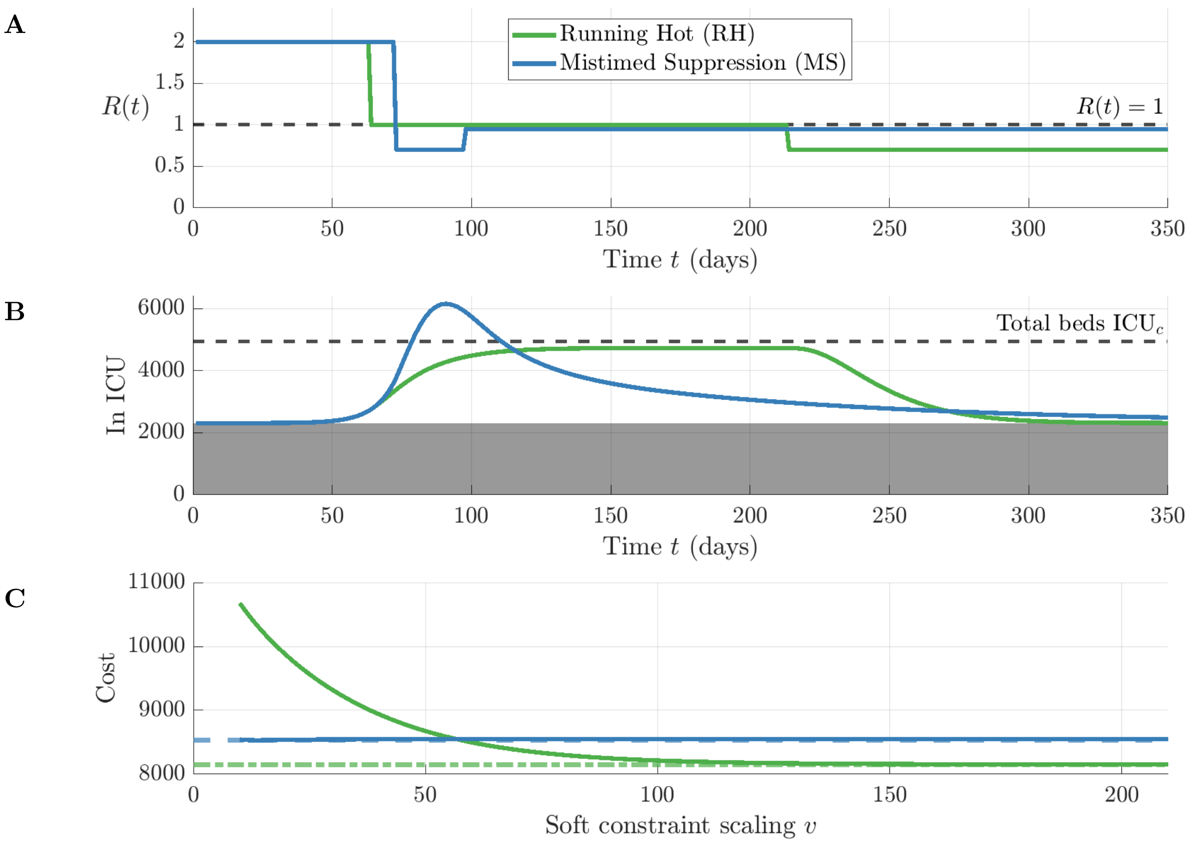


**Figure S11:** The simulation of the Running Hot strategy (RH, green) alongside the Mistimed Suppression strategy (MS, blue) and their costs under the soft constraint and $\alpha_1 = 2\alpha_0 = 0.6$. Each strategy is simulated from a time series for the reproduction number $R(t)$ (A), leading to drastically different dynamics (B). Here, the timing of the $R(t) = 0.7$ intervention for Strategy MS is one day later (day 73) in comparison to the main text (day 72). The dashed black line indicates capacity to treat disease patients $(1 - \eta^{\text{ICU}})\text{ICU}_c$. The optimal strategy (defined by lower cost) is dependent upon parameters $v$ scaling the severity of the soft constraint (C). Note that cost is increasing on the y-axis and so the optimal strategy (defined by lower cost) lies below its counterpart.

#### 7 Sensitivity to the timing and effectiveness of interventions

Here we demonstrate robustness of results to changes in the intervention specifications of Strategies ES and MS for scenarios 1 and 2 respectively. In the first scenario, we kept the $t, R(t)$ specification of Strategy LS from the main analysis ($R(t) = 2$ for times $t \in [1, 70)$ and $R(t) = 0.7$ for times $t \geq 70$). We then allowed the intervention implemented in Strategy ES to vary with some beginning time $t_x \in \{60, 61, \ldots, 70\}$ and its resulting efficacy to vary, $R(t) \in [0.65, 1]$ for $t \geq t_x$. There were a range of $t_x, R(t \geq t_x)$ values where Strategy ES was optimal under the soft constraint but sub-optimal under the hard constraint (Fig S12, light orange). The earlier the intervention was implemented (lower $t_x$), the less effective it needed to be at suppressing transmission for the strategy to be deemed optimal over Strategy LS. The soft constraint relaxed this requirement so that the post-intervention $R(t \geq t_x)$ required for optimality was greater in comparison to the hard constraint (comparing the dark and light orange in Fig S12).

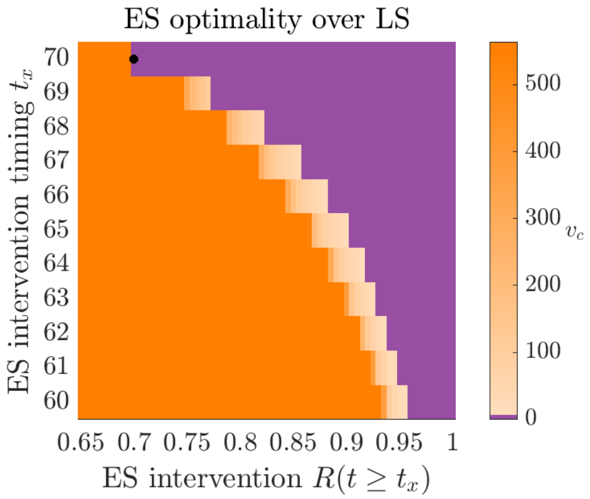


**Figure S12:** Strategy ES optimality (defined as lower cost) over Strategy LS under various scenarios which alter the timing $t_x$ and post-intervention $R(t \geq t_x)$. Strategy LS is fixed with the same $t, R(t)$ specification as the main text (highlighted with black circle). Dark orange represents the region where ES is optimal under both the hard and soft constraint, purple represents the region where ES is sub-optimal under both the hard and soft constraint, and light orange indicates scenarios where ES is sub-optimal under the hard constraint, but optimal under the soft constraint for values $v < v_c$.

Next, we investigated the second scenario where we maintained the $t, R(t)$ specification of Strategy RH from the main analysis ($R(t) = 2$ for times $t \in [1, 64)$, $R(t) = 1$ for times $t \in [64, 214)$, and $R(t) = 0.7$ for times $t \geq 214$). We varied the timing and efficacy of the intervention implemented by Strategy MS ($t_x \in \{65, 66, \ldots, 75\}$, $R(t) \in [0.5, 1]$ for $t \in [t_x, t_x + 25)$, and $R(t \geq t_x + 25) = 0.95$). We obtained a similar result to above where delays in intervention timing $t_x$ resulted in a range of post-intervention $R(t), t \in [t_x, t_x + 25)$ values where Strategy MS was optimal under the soft constraint but sub-optimal under the hard constraint (Fig S13, light blue). This was true even when the peak ICU occupancy under Strategy MS still exceeded the number of beds available (the red marks in Figure S13). The later the intervention under Strategy MS was implemented (higher $t_x$), the more effective it needed to be at reducing transmission to ensure optimality over Strategy RH. Again, the soft constraint required a higher effective $R(t)$ for optimality to occur given a delay in intervention timing (comparing the dark and light blue in Fig S13).

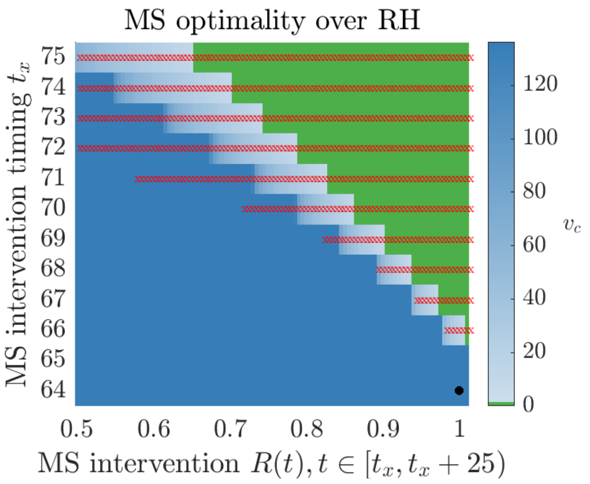


**Figure S13:** Strategy MS optimality (defined as lower cost) over Strategy RH under various scenarios which alter the timing $t_x$ and post-intervention $R(t \geq t_x)$. Strategy RH is fixed with the same $t, R(t)$ specification as the main text (highlighted with black circle). Dark blue represents the region where MS is optimal under both the hard and soft constraint, green represents the region where MS is sub-optimal under both the hard and soft constraint, and light blue indicates scenarios where MS is sub-optimal under the hard constraint, but optimal under the soft constraint for values $v < v_c$. A red x is used to mark the scenarios where the peak ICU occupancy under Strategy MS exceeds the number of beds available.

#### 8 Results with discounting costs at 1.5% per annum

In this section we present our main results where the cost function is adapted to incorporate discounting. This is a common practice in the health economics literature whereby policy makers are assumed to assign higher weighting to imminent costs over long-term costs. The United Kingdom appraises health economic policies with risk to health and life at a discount rate of 1.5% per annum [6]. Thus, our cost function for either health constraint becomes,

$$\sum_{t=1}^{\infty} \gamma(t)\mathrm{ICU}(t) \sum_{s=t}^{\infty} \alpha(s)\bar{T}^{H}(s-t), \tag{6}$$

where

$$\gamma(t) = \frac{1}{(1+0.015)^{t/365}}. \tag{7}$$

Fig S14 shows the costs of the Running Hot (RH) and Mistimed Suppression (MS) control strategies in the presence and absence of discounting. The change in threshold $v$ for which the optimal strategy switches changes only marginally (54.47 without discounting compared to 54.18 with discounting). Since our control strategies are simulated over a relatively short ($< 3$ year) time horizon, discounting this has a negligible effect on strategy costs and our key results are robust to the inclusion or exclusion of discounting.

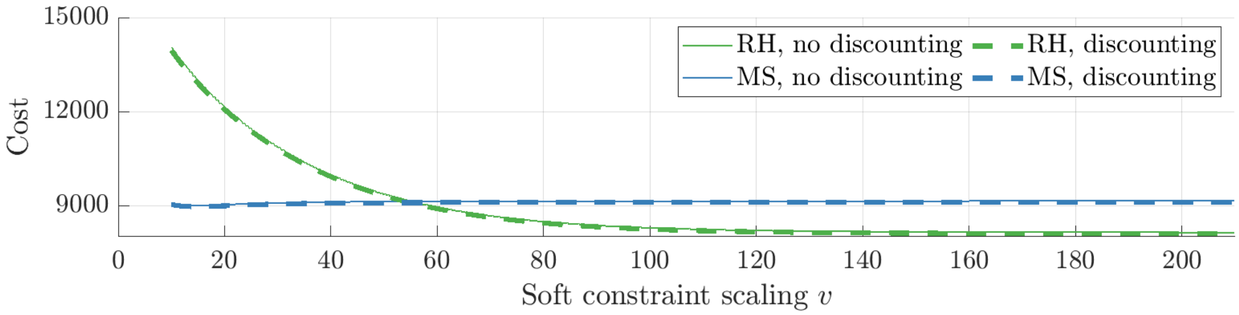


**Figure S14:** Costs of the Running Hot (RH, green) and the Mistimed Suppression (MS, blue) control strategies in the presence of discounting (dashed lines) in comparison to no discounting (solid lines)
